# Risk factors influencing local cholera transmission in urban and rural endemic zones: the experience of Goma and Bukama, Democratic Republic of the Congo, 2021-2022.

**DOI:** 10.64898/2026.07.23.26358563

**Authors:** Kabier Izzeldin, Eve Rahbé, Rachel Mahamba, Hawa Yaffa, Veselina Yosifova, Daniel Mukadi-Bamuleka, Jacques Muzinga, Espérance Tsiwedi-Tsilabia, Faida Kitoga, Sophie Meakin, Tavia Bodisa-Matamu, Flavio Finger, Deka Kabunga, Toto Kyungu, Marie-Laure Quilici, Placide Okitayemba-Welo, Anaïs Broban

## Abstract

While cholera imposes a substantial burden across the Democratic Republic of the Congo, its transmission dynamics vary by geography, socio-economic context or population habits. We investigated risk factors associated with laboratory-confirmed cholera among suspected cases in urban Goma and rural Bukama sites, across 2021-2022. Mixed-effects logistic regression models were fitted independently by site. Positivity rates were higher in Goma (70.2%, 1688/2405) than in Bukama (49.4%, 390/790). In both sites, rainy seasons increased odds of testing positive (OR=3.89, 95%CI [2.93-5.16] and 2.00 [1.39-2.86] for Goma and Bukama, respectively) while receiving ≥1 Oral Cholera Vaccine dose was protective (0.53 [0.37-0.74] and 0.58 [0.37-0.91]). Some risk factors were significant only in Goma, including displacement status (1.93 [1.11-3.35]), household cholera case exposure (1.79 [1.31-2.45]), consumption of surface water (2.11 (1.46-3.03]) or from water tankers (1.75 (1.24-2.47]). This study shows that cholera risk factors vary across endemic settings, giving insights for improved, locally-adapted control measures.

## Introduction

Cholera is an acute diarrheal disease caused by toxigenic strains of the bacterium *Vibrio cholerae* of the O1, and more rarely O139 serogroups, whose clinical presentation includes abdominal cramps, profuse watery diarrhea, and emesis (1). It remains a major global health threat, with an estimated 1.3 to 4.0 million cases annually worldwide (2). Following a period of relative decline, reported global cholera incidence and mortality have resurged in recent years (3,4) with sub-Saharan Africa continuing to bear the greatest share (5).

The Democratic Republic of the Congo (DRC) has consistently ranked among the most affected countries and is currently facing the highest number of cholera cases and deaths in the African region. In 2024, the country reported over 31,000 suspected cholera cases and 400 associated deaths (4,6). The eastern part of DRC has historically been the most affected due to a complex history of conflicts and humanitarian emergencies, perpetuated by mass displacements, fragile health systems, food insecurity, and poor access to water, sanitation and hygiene (WASH). Cholera became notably endemic in the Kivu provinces, in part following the 1994 influx of approximately 800,000 Rwandan refugees in the aftermath of the genocide. Goma city has since experienced several outbreaks of importance, the last of which began in 2022 following the arrival of displaced people from rural North Kivu areas (7). Cholera has also taken root in the broader African Great Lake regions, such as Tanganyika or Haut-Lomami provinces (6,8).

Cholera generally manifests at the intersection of systemic inequities in WASH infrastructure, seasonal weather patterns, weak health systems and socioeconomic status. Its transmission is further exacerbated in conflict-afflicted regions, where forced displacement renders the population particularly vulnerable (9,10). Individual-level transmission mainly happens through the consumption of contaminated water and food, but may also occur via human-to-human contact and fomites (11,12).

Understanding the local transmission patterns in cholera-affected and endemic zones is crucial for implementing effective measures to ensure cholera control in the region. Nevertheless, characterizing local cholera transmission remains subject to recurrent constraints, such as non-specific case definition and difficulties with systematic laboratory case confirmation and investigation (13).

The present study aimed to characterize local cholera risk and protective factors in two endemic sites, one urban (Goma, North Kivu) and one rural (Bukama, Haut Lomami). The association of socio-demographic, climate and behavioral factors with laboratory-confirmed cholera among suspected cases presenting to Cholera Treatment Units (CTUs) in the two study sites are assessed, for a period ranging from May 2021 to December 2022.

## Methods

### Overall project

The present study is part of a multi-center research project in the DRC implemented since 2021 by Epicentre and its local partners, aiming at evaluating the impact of the Oral Cholera Vaccine (OCV) preventive strategy. The last 2-doses campaigns before or during the study period happened in May/October 2019 and January/June 2020 in Goma, and December/April 2021-2022 in Bukama. One of the main activities of the project is reinforced clinical surveillance with laboratory confirmation of suspected cholera cases and routine data collection in both Goma city in the North-Kivu Province and Bukama, a rural health zone in the Haut-Lomami Province.

### Study design and participants

The study included all active CTUs across the two study sites during 2021 and 2022 (Figure 1). In Goma, situated on the northern shore of Lake Kivu, 11 CTUs were drawn from four health zones (Goma, Karisimbi, Kirotshe, and Nyiragongo) spanning the urban and peri-urban areas of the city, and the closest agglomeration (Saké). In Bukama, 9 CTUs from one health zone, which borders the Lualaba river (upper course of the Congo River) in the ex-Katanga region, were included.

**Figure 1.**
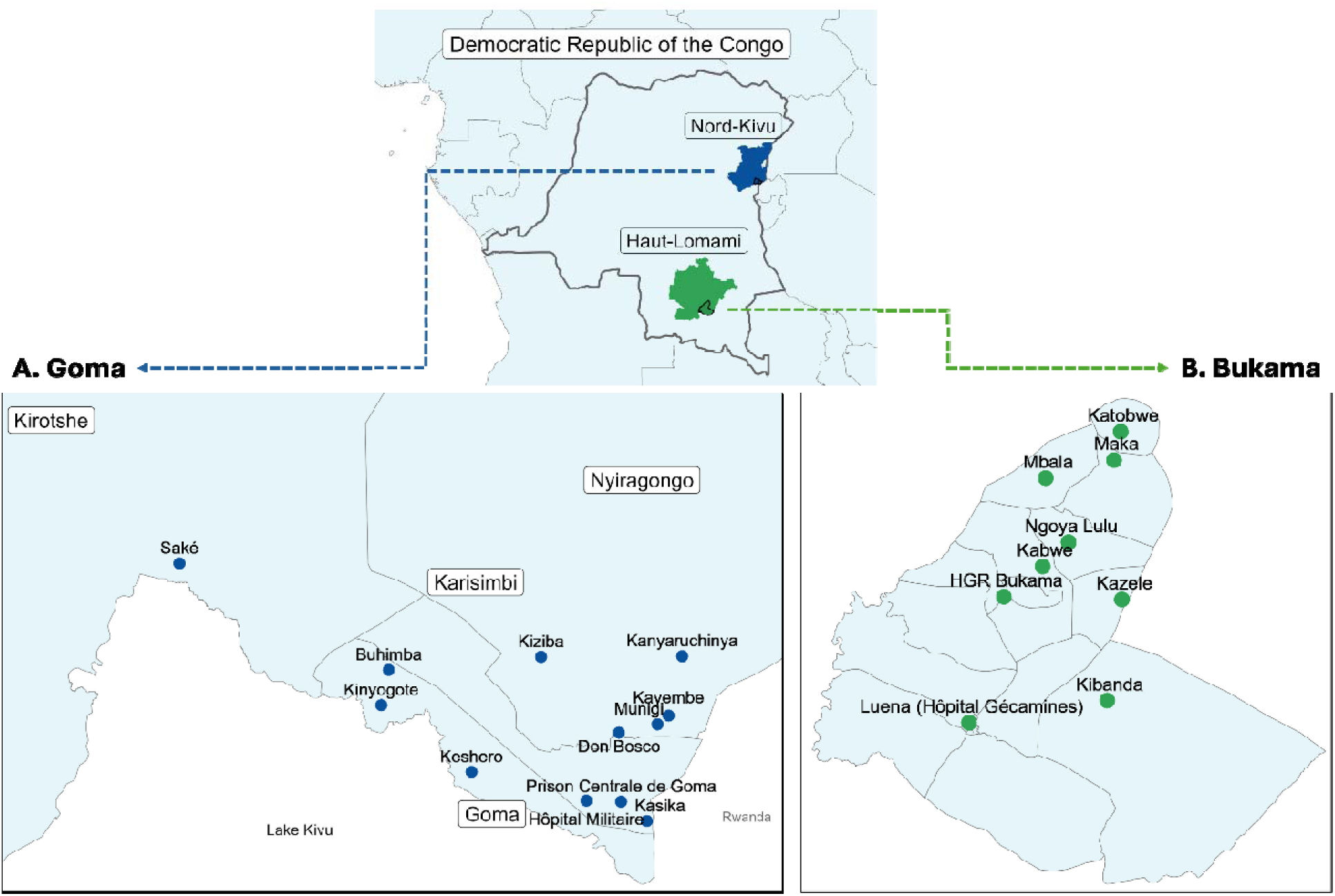
Map of local health zones/areas and Cholera Treatment Units (CTUs) included in the study across Goma, Nord-Kivu province (blue, **A. panel**) and Bukama, Haut-Lomami province (green, **B. panel**), within the Democratic Republic of the Congo.

All individuals presenting to participating CTUs with suspected cholera between May 12, 2021, and December 31, 2022, in Goma, and between October 17, 2021, and December 31, 2022, in Bukama, were eligible to participate in the study. Suspected cholera was defined as a patient reporting more than three watery stools within 24 hours, in accordance with national guidelines (14).

### Case management and data collection

Epicentre-supported Ministry of Health (MoH) nurses were appointed to CTUs managed by either the MoH, MSF, or partner non-governmental organizations (NGO). Their training included general cholera knowledge, study design, informed consent procedures, patient interviews, and collection, packaging and shipment of biological samples.

Nurses administered paper-based questionnaires and collected fresh stool samples at the CTU-level from consenting study participants. The same level of care was applied to all patients, according to local CTU standards, irrespective of their participation in the study or any test results.

Both wet and dry filter papers were prepared from fresh stool samples in accordance with the Global Task Force on Cholera Control (GTFCC) guidance (15). Questionnaires investigated multiple factors, all self-reported: sociodemographic (sex, age, displacement status), OCV status (i.e. number of doses received; dates), household information (household education level, number of household members), behavioural (hygiene practices – handwashing method and habits; main source of drinking water – public/private tap, surface water, rainwater, water tanker, unprotected sources, borehole), sanitation (toilet infrastructure – flush tank, pit with/without slab, hanging toilets over lake/river, open defecation), and environmental (seasonality, exposure to cholera cases) (Supp. S1). Participants could report up to three drinking water sources and latrine types regularly used. All data were pseudonymized at the CTU level.

### Laboratory procedures

#### Culture

Wet filter paper samples were shipped by road to Rodolphe Mérieux Institut National de Recherche Biomédicale (INRB) Goma for Goma samples, and in Grand Laboratoire of Lubumbashi (GLL) for Bukama samples. Samples were enriched in alkaline peptone water (APW, Oxoid Thermo Scientific ™) and then grown on TCBS medium (Oxoid Thermo Scientific ™). Suspected bacterial colonies were isolated on a non-selective TSA medium (Oxoid Thermo Scientific™) and exposed to polyvalent *V. cholerae* antiserum (MAST diagnostic), in accordance with cholera surveillance guidelines (15).

#### PCR

Samples were sent by air to the Institut Pasteur in Paris and handled in compliance with IATA regulations. DNA was extracted from dried filter paper samples through heat-induced cell lysis. Additional products resulting from cell lysis were absorbed with BT Chelex® resin in accordance with previously published methods (16,17).

Samples were tested at the Pasteur Institute in Paris and at INRB Goma either through conventional PCR against the *O1rfb* gene (18) or through a Research Use Only product triplex qPCR targeting the *toxR* species-specific, *O1rfb* serogroup-specific, and *ctxA* toxin-specific genes. In each qPCR well, a 5μL sample of DNA was added to a 15μL mixture of primer, probe, molecular grade water, and a Taqman multiplex with Mustang Purple reference dye (ThermoFisher, USA). Standard curves of well-characterized *V. cholerae* O1 and two negative controls were added to each plate. Assays were then processed with a QuantStudio 5 thermocycler with the following conditions: 20-second hold at 95°C, 40 3-second cycles of 95°C, and a 30-second hold at 61°C. Results analysis was conducted utilizing the Design & Analysis Software 2.6.0 (ThermoFisher, USA) and validated manually using calculated thresholds and curves. Patient samples with qPCR testing were considered positive for cholera if *O1rfb* detection after amplification was reached under 37 cycles.

## Statistical Analysis

Final cholera positivity status was determined for each sample according to the criteria in Box 1. Analyses for Goma and Bukama were conducted separately to reflect local epidemiological contexts. All analyses and data visualization were conducted using R version 4.4.3.

**Suspected cholera case definition:** Patient reporting more than three watery stools within 24 hours.

**Laboratory-confirmed cholera case:**

- **Positive:**

`o` Detection of the *O1rfb* gene by qPCR or conventional PCR, OR
`o` Isolation of *Vibrio cholerae* belonging to the O1 serogroup by culture.
- **Negative:**

`o` Negative results by both PCR and culture tests, OR
`o` Negative PCR result only when culture was not performed or was inconclusive.

Box 1. Criteria for classification of both suspected and laboratory-confirmed cholera cases, Democratic Republic of the Congo, May 2021-December 2022.

Ten imputed datasets were computed using multiple imputation with chained equations for multivariate modelling purposes, addressing missing values observed in predictor variables (Supp. S2 Figure 1,2) (19,20). Imputation diagnostics were further conducted using data visualizations (i.e., mean diagnostic plots, kernel density plots, box plots, and residual plots) (Supp. S2 Figure 3, 4) (21).

**Figure 2.**
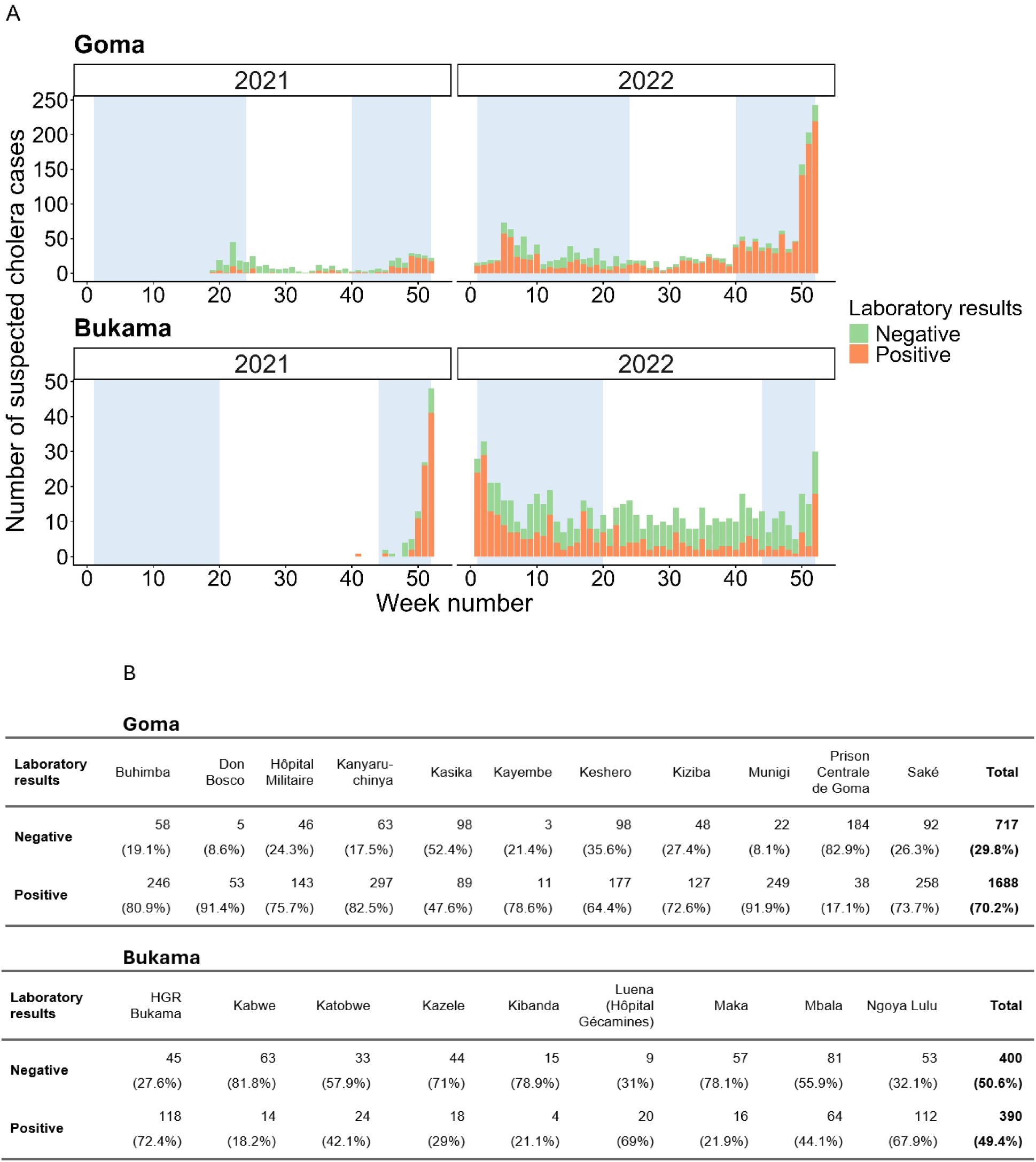
**A.** Epidemiological weekly curve of suspected cholera cases presenting to the CTUs included in the surveillance study with associated laboratory results, by year and study site. Blue zones represent rainy seasons: September to May for Goma, and October to April for Bukama. **B.** Number and positivity/negativity rates of suspected cholera cases by Cholera Treatment Units (CTUs).

Univariate analyses were first conducted using simple logistic regression, with cholera positivity as the outcome variable. Variables to be tested in the multivariate models were selected using an associated p-value threshold of α=0.2 (Supp. S4). Multivariate analyses were conducted using mixed-effects logistic regression, with a random intercept at the CTU level to account for site-specific clustering, and included sex and age group. Other variables were selected following the stepwise backward-forward model-building approach (22), using a p-value threshold of α=0.2. Pooled Odds Ratios (ORs) across the ten imputed datasets were computed according to Rubin’s rule (23), and 95% confidence intervals (95%CI), and p-values were generated. A significance threshold of α=0.05 was considered. A generalized variance inflation factor was computed to test multicollinearity among final model variables (24). Supp. S2 Table 3 shows results from the multivariate model on non-imputed datasets.

## Ethics Approval

This study received ethical approval by Médecins Sans Frontières Ethics Review Board (MSFERB n°2104) and Comité National d’Ethique de la Santé (CNES) in Kinshasa, DRC (initial approval number: 248/CNES/BN/PMMF/2020). This study adhered to the Council for International Organizations of Medical Sciences (CIOMS) International Ethical Guidelines for Health-related Research Involving Humans (25). Oral informed consent was obtained from all participants, or from their guardians for minor participants; oral assent was collected for participants aged 13-17 years. Consents and assents were documented. Participation in the study was voluntary.

## Results

### Description of participants

Of the 2504 participants from Goma and 800 from Bukama initially included in the clinical surveillance in 2021-2022, a total of 2405 (96%) and 790 (99%) had associated laboratory results for their cholera statuses and were included in the final analysis, for Goma and Bukama respectively.

Figure 2.A shows the weekly curve of suspected cholera cases presenting at the two study sites and Supp. S3 additionally shows monthly positivity rates and laboratory results by study sites and CTUs. In Goma, positive cholera cases were mostly reported from Kanyaruchinya, Saké, Munigi and Buhimba CTUs – representing population coming from health areas where camps were installed by the end of 2022 (Figure 2.B.). In the rural site, the highest positive case numbers were from the CTUs of the General Referral Hospital (HGR) in Bukama center and of Ngoya Lulu (Figure 2.B.). There was a median of 222 [IQR: 181-289.5] participants per CTU in Goma and 73 [IQR: 57-145] in Bukama.

Overall positivity rate was 70.2% (1688/2405) in Goma and 49.4% (390/790) in Bukama (Figure 2.B. and Table 1). Displaced people were exclusively reported in Goma by the end of 2022, 91% of whom were positive for cholera. The proportion of individuals who received at least one OCV dose differed substantially between Goma (10.6%, 254/2405) and Bukama (79.9%, 631/790). The mean patient age was 16.7 years in Goma and 23.6 years in Bukama, with substantially different age distributions among suspected cholera cases. Positivity rates were highest during rainy seasons in both Goma and Bukama but remained substantial during the dry seasons (45% and 36% for Goma and Bukama respectively).

**Table 1.** Main socio-demographic variables and associated positivity rates to cholera in the population of study, for both sites in the Democratic Republic of the Congo, 2021– 2022.

| Characteristics | Goma |  |  | Bukama |  |  |
| --- | --- | --- | --- | --- | --- | --- |
|  | Negative N = 717<sup>1</sup> | Positive N = 1,688<sup>1</sup> | Overall N = 2,405<sup>1</sup> | Negative N = 400<sup>1</sup> | Positive N = 390<sup>1</sup> | Overall N = 790<sup>1</sup> |
|  | <sup>1</sup>n (%): cholera positivity rates |  |  |  |  |  |
| **Year** |  |  |  |  |  |  |
| 2021 | 265 (62%) | 164 (38%) | 429 (100%) | 19 (19%) | 82 (81%) | 101 (100%) |
| 2022 | 452 (23%) | 1,524 (77%) | 1,976 (100%) | 381 (55%) | 308 (45%) | 689 (100%) |
| **Age** |  |  |  |  |  |  |
| <5 | 204 (32%) | 436 (68%) | 640 (100%) | 37 (48%) | 40 (52%) | 77 (100%) |
| 5-14 | 75 (11%) | 631 (89%) | 706 (100%) | 68 (46%) | 81 (54%) | 149 (100%) |
| 15-24 | 165 (39%) | 253 (61%) | 418 (100%) | 130 (57%) | 98 (43%) | 228 (100%) |
| 25-44 | 193 (41%) | 283 (59%) | 476 (100%) | 122 (49%) | 129 (51%) | 251 (100%) |
| 45+ | 80 (48%) | 85 (52%) | 165 (100%) | 43 (51%) | 42 (49%) | 85 (100%) |
| **Sex** |  |  |  |  |  |  |
| Female | 270 (26%) | 757 (74%) | 1,027 (100%) | 195 (51%) | 185 (49%) | 380 (100%) |
| Male | 447 (32%) | 931 (68%) | 1,378 (100%) | 204 (50%) | 205 (50%) | 409 (100%) |
| (Missing) |  |  |  | 1 | 0 | 1 |
| **≥1 dose OCV** |  |  |  |  |  |  |
| No | 580 (27%) | 1,531 (73%) | 2,111 (100%) | 52 (33%) | 104 (67%) | 156 (100%) |
| Yes | 128 (50%) | 126 (50%) | 254 (100%) | 347 (55%) | 284 (45%) | 631 (100%) |
| (Missing) | 9 | 31 | 40 | 1 | 2 | 3 |
| **Displacement Status** |  |  |  |  |  |  |
| No | 668 (36%) | 1,202 (64%) | 1,870 (100%) | 400 (51%) | 390 (49%) | 790 (100%) |
| Yes | 49 (9%) | 486 (91%) | 535 (100%) | 0 | 0 | 0 |
| **Season** |  |  |  |  |  |  |
| Dry | 210 (55%) | 169 (45%) | 379 (100%) | 155 (64%) | 88 (36%) | 243 (100%) |
| Rainy | 507 (25%) | 1519 (75%) | 2,026 (100%) | 245 (45%) | 302 (55%) | 547 (100%) |

### Cholera risk factors in Goma

Variables retained in the final multivariate mixed-effect logistic regression model for Goma are presented in Table 2. Unadjusted estimates from univariate simple logistic regression models, used for variable selection, are shown in Supp. S4. The age group most at risk of testing positive for cholera in Goma were the 5-14 years old children, with 3 times the odds compared to 15-24-year-old young adults. Then, the odds of testing positive for cholera decreased significantly with age compared to young adults. The issue of displacement was prominent across Goma, where displaced individuals carried almost 2 times the odds of cholera positivity compared to non-displaced individuals (OR=1.93 with 95% confidence intervals [1.11-3.35]). Exposure to a suspected cholera case living in the same household increased the odds of cholera positivity by 79% (OR=1.79 [1.31-2.45]). Participants with at least one OCV dose had 47% decreased odds of cholera positivity compared to unvaccinated individuals (OR=0.53 [0.37-0.74]). Rainy seasons in Goma (September to May) were also strongly associated with increased cholera positivity compared to dry periods (OR=3.89 [2.93-5.16]). Associations with sex, education level and handwashing method were not significant in the final multivariate model. Water tankers, which draw water primarily from lake surface water, were the most reported primary drinking water sources amongst participants (46%, 1090/2405, Supp. S4). Water tanker and surface water sources were observed to increase individuals’ odds of cholera positivity (OR=1.75 [1.24-2.47] and 2.11 [1.46-3.03] respectively) whereas rainwater was a non-significant protective factor. Moreover, the use of tank-based flush toilets was estimated to be a significant protective factor (OR=0.37 [0.20-0.69]).

**Table 2.**
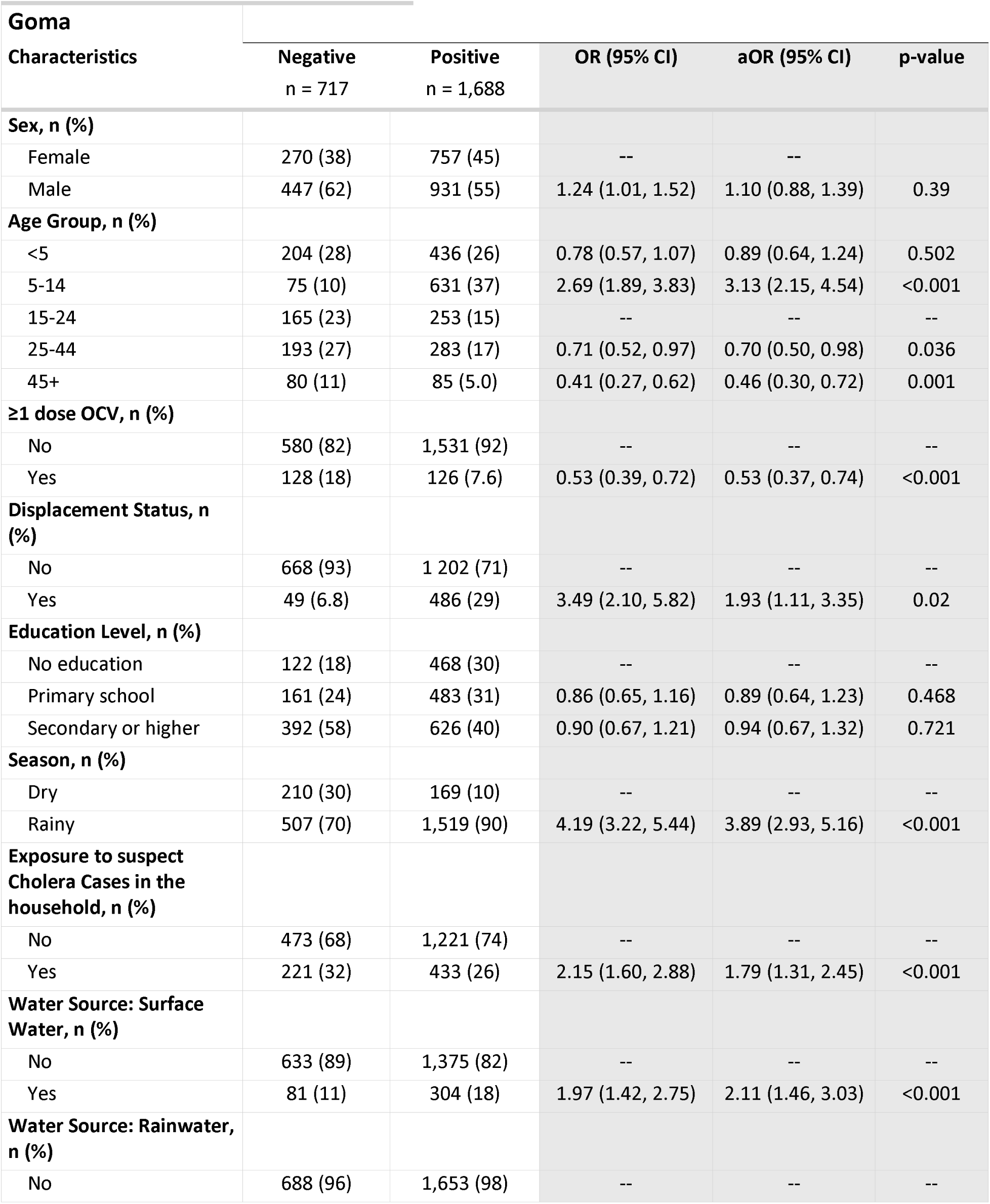
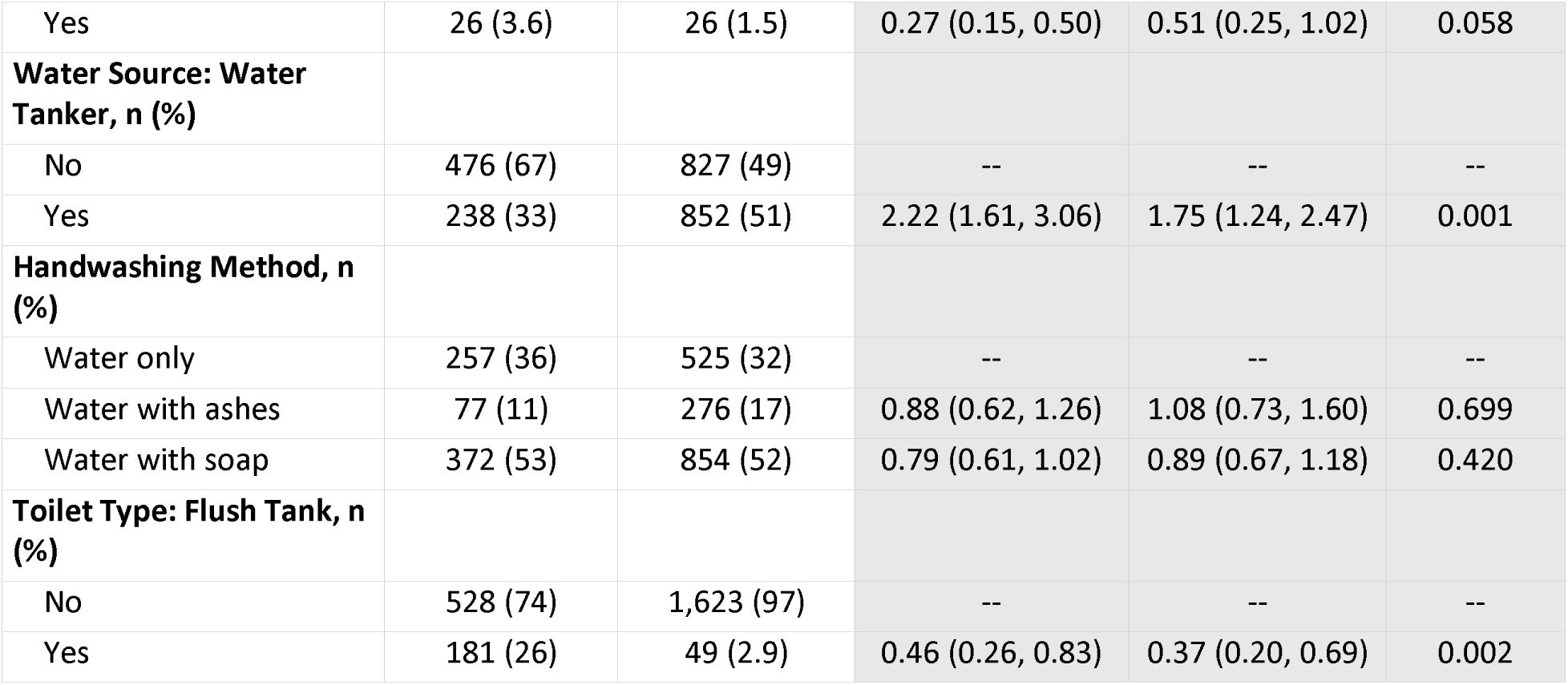
Multivariate mixed-effect logistic regression model outputs for laboratory-confirmed cholera positivity among suspected cases with Odds Ratio (OR, univariate results) and adjusted Odds Ratios (aOR, multivariate results), 95% Confidence Intervals and p-values, in Goma, Democratic Republic of the Congo, 2021–2022.

**Table 3.**
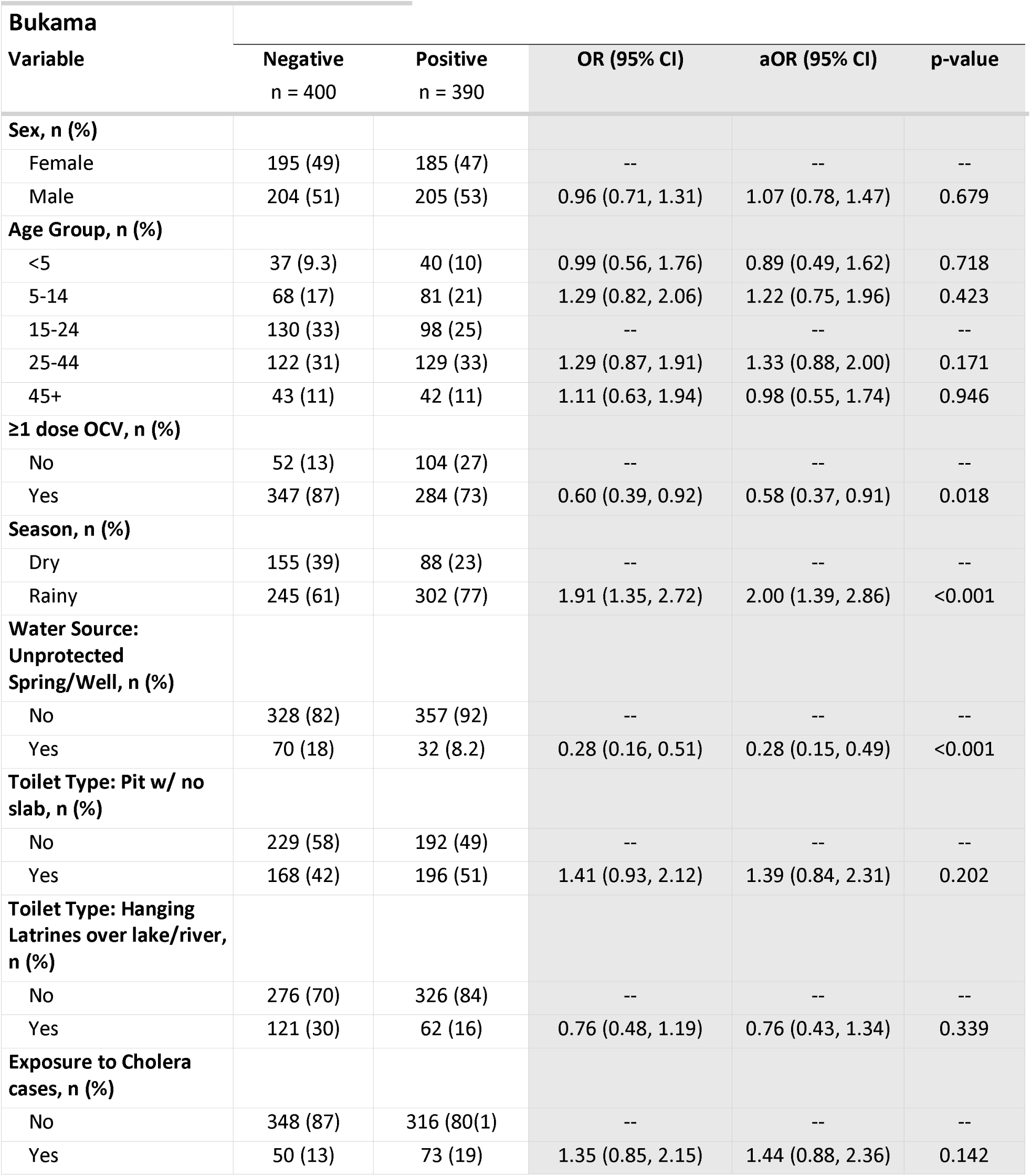
Multivariate mixed-effect logistic regression model outputs for laboratory-confirmed cholera positivity among suspected cases with Odds Ratio (OR, univariate results) and adjusted Odds Ratios (aOR, multivariate results), 95% Confidence Intervals and p-values, in Bukama, Democratic Republic of the Congo, 2021–2022.

### Cholera risk factors in Bukama

Variables retained in the final multivariate mixed-effect logistic regression model for Bukama are presented in Table 2. Unadjusted estimates from univariate simple logistic regression models, used for variable selection, are shown in Supp. S4. In contrast to Goma, there was no clear association between positivity to cholera and age. There was no cholera-positivity association with sex either. Participants with at least one OCV dose had 42% decreased odds of cholera positivity compared to unvaccinated individuals (OR=0.58 [0.37-0.91]). Rainy seasons in Bukama (October to April) doubled the odds of testing positive for cholera compared to dry periods (OR=2.00 [1.39-2.86]). Exposure to suspected cholera cases in the household and use of latrine pit without slab were non-significant risk factors, whereas using hanging toilets over lake/river was a non-significant protective factor.

Regarding water sources, the use of unprotected springs or wells unexpectedly reduced the odds of cholera positivity by 72% (OR=0.28 [0.15-0.49]).

However, the distribution of water sources across CTUs (representing different catchment areas and populations), varied substantially—particularly between the semi-urban CTU of HGR Bukama and the other CTUs. To address these differences, we provide a sensitivity analysis (Supp. S5) separating HGR Bukama CTU data from the others and fitting two independent models.

At HGR Bukama, public tap water was almost exclusively reported by patients and was the most frequent source (96%, 156/163), whereas surface water was the most frequent source in all other CTUs (82%, 517/627). At HGR Bukama, 28% drank from unprotected springs or wells, while diversifying their water sources: 15% of them also drank surface water and 89% also drank public tap water (Table S5.2). In contrast, only 3% of users elsewhere diversified their water sources (Table S5.4).

The results of the two models showed that the use of unprotected springs or wells significantly decreased the odds of testing positive for cholera only in HGR Bukama. Thus, patients drinking water from unprotected sources were at lower risk to test positive for cholera compared to those who did not drink it or drank only surface water or public tap water – a significant risk factor in the univariate simple logistic regression.

## Discussion

While general cholera risk factors linked to water, social and behavioural factors are widely established (26), our study sheds light on these individual risk factors in two endemic yet very different sites of a heavily cholera-affected country, the Democratic Republic of the Congo (DRC). Overall, our findings show how cholera infections are influenced by local characteristics while confirming OCV protects individuals, and give context-specific operational insights for limiting its burden.,n addition, we have explored cholera risk factors for laboratory-confirmed infections among suspected cases, which was rarely done before, highlighting specific determinants of transmission and leverages to control cholera over other diarrheal diseases.

Our results first showed common risk and protective factors across cholera endemic sites. Findings confirmed that cholera risk is seasonal: rainy seasons were estimated to be a significant risk factor for individual cholera infection in both Goma and Bukama. Nevertheless, positivity rates indicated that infections also occurred during dry seasons, suggesting that cholera control should not be neglected during this period. In both study contexts, and despite intermediate vaccine coverage levels in Goma (27) or differences in the participants’ socio-demographic between the two sites (Table 1), being vaccinated with at least one dose against *Vibrio cholerae* provided significant individual protection. These findings are consistent with vaccine effectiveness estimates in similar contexts (28,29).

Results also highlighted specific risk factors for an urban site such as the Goma city area. The presence of another case of acute watery diarrhea in the same household significantly increased the odds of being infected with cholera in Goma, consistent with previous studies and with cholera generally occurring in clusters, including in endemic areas (30,31). This highlights the importance of within-household transmission, especially in dense urban areas. It also reinforces the strategic value of implementing cholera response at the community level, within households and in neighboring ones, for example through Case-Area Targeted Interventions (CATI) (32). Additionally, while displacement events are known to favor cholera outbreaks (33,34), it was a clear contributing factor to cholera positivity risk in Goma. Massive arrivals of internally displaced people (IDP) starting in October 2022 were followed by subsequent cholera outbreaks, suggesting the introduction of a non-or less-immunized population into this heavily endemic context. Importantly, this massive influx of IDP in Goma coincided with the rainy season at the end of 2022, which might have exacerbated the effect of seasonality in our model.

At the individual level, the two study sites presented different age distributions within suspected cholera cases, with more children presenting in Goma. There was no association with age group in Bukama. In Goma, under-five patients carried lower odds of testing cholera-positive than older children, confirming previous findings and reflecting the more diverse etiology for acute watery diarrhea in this age group (35). Nevertheless, there is a significant trend with older patients having lower odds of testing positive for cholera among acute watery diarrhea patients, which could suggest different endemicity stages, with Goma’s older population able to maintain immunity through possible multiple exposures to cholera over the years.

The importance of WASH-related exposures to cholera is well documented, and improved sanitation facilities are proven to significantly reduce cholera transmission (36,37). Our findings also showed that cholera positivity is associated with hygiene, latrine types or water sources and highlighted the importance of investing in sanitation infrastructure at both community and individual levels. In Goma, individuals who used surface water and water tankers as primary drinking sources were at a greater risk for cholera positivity compared with the average primary water source there. In the DRC, recurring outbreaks were historically linked to lake regions (8,38). The specific geographical configuration of Goma indeed creates a favorable environment for *V. cholerae* circulation and subsequent exposure: presence of volcanic soil impeding soil digging leading to limited access to water sources other than the lake (39), combined with discharge of wastewater nearby drinking sources in the lake, high population density and inadequate water treatment. While water tankers can serve as an improved water source when established correctly, they require systematic water treatment and chlorination, as well as support to vendors in implementing such measures (40).

In the rural context of Bukama, despite the presence of lakes and rivers in the region, the use of surface water did not appear as a prominent risk factor. Lower population density, and subsequent lower close contacts and lower levels of wastewater discharge nearby drinking water collection points, may decrease the odds of contamination through surface water. The observed significant protection of unprotected water sources for cholera positivity was mostly driven by people presenting to the CTU of semi-urban Bukama city (HGR Bukama), where the frequency of public tap water users – the main water distribution infrastructure provided to the population – was high. As unprotected source users also systematically drank from other sources, this might suggest that diversifying water sources beyond the public tap decreases odds of testing positive for cholera. Together with the high odds of cholera associated with consuming public tap water in univariate models (Supp. S4), this points towards public tap water being a potential source of infection in Bukama, possibly due to damaged infrastructure or lack of chlorination. Overall, these findings call for action to improve public infrastructure and water quality to avoid risks of water distribution systems that favor infectious diseases contamination (41).

This study is limited by the self-reported nature of risk factor questionnaires, which could introduce social desirability and memory bias especially regarding vaccination status or hygiene practices. Moreover, differences in risk factors prevalence across CTUs could have influenced our estimates despite the inclusion of a random intercept to account for such differences. In Bukama, sample sizes in certain groups may also have limited statistical power, which may have prevented the formal identification of some predictors. The major strength of this study lies in the use of cholera laboratory results as opposed to non-specific suspected cholera case definitions, which allows to differentiate risk factors for cholera from other diarrheal diseases. In particular, the use of PCR provides higher reliability for cholera confirmation. Nevertheless, risk factors for laboratory-confirmed cholera among suspected cases could differ compared to cases found in the general population, and should be interpreted as such.

Overall, this study provides a detailed understanding of cholera risk and protective factors in two distinct cholera-endemic contexts in the DRC. Despite general knowledge on cholera epidemiology, local geography, infrastructure, and population habits remain key factors shaping cholera circulation. The findings from our study reaffirm the GFTCC’s call for multisectoral action in cholera control, establishing the importance of preventive measures. These include combining long-term WASH improvements, access to appropriate care, community engagement, and preventive OCV campaigns. Notably, WASH interventions should not only emphasize stricter enforcement of safe water distribution regulations and quality controls by local authorities but also promote local behavioral shifts driven by civil society organizations. Furthermore, despite challenges in implementing campaigns in urban settings [25], preventive OCV campaigns in endemic areas appear to be a promising strategic approach, though their real impact needs to be investigated further.

## Supplementary Information

**Additional file S1:** Description of variables used for analysis.

**Additional file S2:** Supplementary information on imputation.

**Additional file S3:** Time and location description of laboratory-confirmed results distribution.

**Additional file S4:** Univariate logistic regression output for laboratory-confirmed cholera positivity with Odds Ratios and 95% Confidence Intervals for Goma and Bukama, Democratic Republic of the Congo, 2021–2022.

**Additional file S5:** Supplementary analysis for the Bukama site.

## Acknowledgement

We are grateful to the surveillance study participants for sharing their information and exposures and accepting to participate in the study. This study would not have been possible without the research teams in the field (especially Patient Kamavu, Joseph Kasereka, Germain Mweha, Nelson Kasongo, Guy Walombe, Nadine Mitutso, and CTU nurses surveyors) and the Médecins Sans Frontières teams in Goma and Lubumbashi. We thank the Ministry of Health of North Kivu and Haut-Lomami, and especially the medical heads of all Health Zones for their support.

## Funding statement

The survey underlying this work was supported by the Wellcome Trust-FCDO grant number: [215689/Z/19/Z]. The funder was not directly involved in the data collection, analysis, or decision to publish these results.

## Authors’ contributions

**Conceptualization and study design:** Anaïs Broban. **Data collection:** Rachel Mahamba. **Supervision:** Placide Okitayemba-Wela, Deka Kabunga, Toto Kyungu. **Administrative, technical or material support:** Rachel Mahamba, Anaïs Broban. **Laboratory support:** Daniel Mukadi, Jacques Muzinga, Tavia Bodisa-Matamu, Espérance Tsiwedi-Tsilabia, Faida Kitoga, Hawa Yaffa, Marie-Laure Quilici, Veselina Yosifova. **Statistical analysis:** Kabier Izzeldin, Eve Rahbé. **Acquisition, analysis or interpretation of data:** Kabier Izzeldin, Anaïs Broban, all authors. **Writing - original draft preparation:** Kabier Izzeldin, Anaïs Broban, Eve Rahbé. **Writing - review and editing:** all authors. Anaïs Broban is the guarantor of the study.

## Competing interests

The authors have no competing interests to declare.

## Data availability

Epicentre is committed to sharing and disseminating health data from its programs and research in an open, timely, and transparent manner to promote health for populations while respecting ethical and legal obligations towards patients, research participants, and their communities. For the purpose of this study, Epicentre collected identifiable participant data.

Upon publication and for as long as the ethical authorisation permits it, the data set underlying the findings of this study will be made available on request. If scientifically relevant, the request may be granted in accordance with legal framework set forth by Epicentre data sharing policy available on its website (statics.teams.cdn.office.net/evergreen-assets/safelinks/2/atp-safelinks.html).

All data access request for non-commercial and academic research can be addressed to or the corresponding authors. Such request will be submitted to the Data Sharing Committee of Epicentre. Data will then be shared upon committee approval and establishment of an appropriate data sharing agreement respecting legal framework, applicable data protection laws, and accounting for the data nature and sensitivity.

## Notes

### Competing Interest Statement

The authors have declared no competing interest.

### Author Declarations

This study received ethical approval by Medecins Sans Frontieres Ethics Review Board (MSFERB n. 2104) and Comite National d Ethique de la Sante (CNES) in Kinshasa, Democratic Republic of the Congo (initial approval number: 248/CNES/BN/PMMF/2020).

