## Supplementary material for "Risk factors influencing local cholera transmission in urban and rural endemic zones: the experience of Goma and Bukama, Democratic Republic of the Congo, 2021-2022.": Supp. S1

**Additional file S1: Description of variables used for analysis** – Tables S1.1 & S1.2

**Table S1.1:** Definition of variables.

| **Variable name** | **Detailed variable definition** | **Method of measurement** | **Variable Class** |
| --- | --- | --- | --- |
| Sex | Sex of participant | Response by participant | Categorical |
| Age | Age of cholera patient stratified by age bracket. Age provided in years and categorized according to the following age brackets:  <5,5-14,15-24,25-44,45+ | Reported by participant and derived by data inspector. | Categorical |
| Year | Year of case inclusion presenting to CTC | Response provided by data collector. | Categorical |
| Vaccination Status | Vaccination status (‘ever vaccinated’) of participant for the OCV during a vaccine campaign in the last 3 years. | Response by participant | Categorical |
| Displacement Status | Participant considers themselves displaced or participant comes from a time period of a CTC that aligns with contextual events (suspected case peaks) for displacement.   1- Individuals answering Yes to “do you consider yourself displaced?” OR Individuals presenting to the following CTCs in Goma at any time period: Bulengo, Don Bosco, Kayembe, Kinyogote, Lushagala, Mugunga, Munigi, Nzulo OR individuals presenting to Buhimba between peak season (March 2023, March 2024, Dec 2024), or Kanyarutshinya (Jan 2023) or Saké (March-April 2023, Nov 2023-Jan 2024).  0- Individuals answering “No” to “do you consider yourself displaced?” or come from one of the following CTCs in goma: Hopital Militaire, Kasika, Keshero, Kiziba, Prison Centrale de Goma OR present to the following CTCs outside of peak suspected case season mentioned above: Buhimba, Kanyarutshinya, Saké | Response by participant and derived by data inspector | Categorical |
| Season | The participants’ date of inclusion to a CTC is from specific months constituting a wet or dry season:   - Goma (Rain): 1, 2, 3, 4, 5, 9, 10, 11, 12 - Goma (Dry): 6, 7, 8 - Bukama (Rain): 1, 2, 3, 4, 10, 11, 12   Bukama (Dry): 5, 6, 7, 8, 9 | Response provided by data collector | Categorical |
| Exposure to suspect Cholera cases in the household | The participant had exposure to other cases of severe diarrhea in their household in the past week prior to presenting to CTC | Response by participant | Categorical |
| **Household information** | | | |
| Household education level | Participant indicated the highest level of education attained by their household/block head | Response by participant, with no education category collapsed from “no education (literate)” and “no education (illiterate) | Categorical |
| Number of children under-5 living in household | The participant indicated a numeric value of the number of children under the age of 5 living in their household. Individuals responding with a numeric value ≥3 were aggregated. | Response by participant and derived by data inspector | Categorical |
| Household size | The participant was asked “how many people currently live in your household ?”   - Small: ≤ 3 individuals in household - Medium: >3 and ≤6 - Large: >6   Institutional: Participants belonging to the following CTCs: "Prison Centrale de Goma", "Hôpital Militaire", "Luena (Hôpital Gécamines)" | Response by participant and derived by data inspector | Categorical |
| **Hygiene practices** | | | |
| Handwashing Method | The participants’ primary method of washing their hands. | Response by participant | Categorical |
| Washes hands before cooking | The participant indicated that they wash their hands before cooking. | Response by participant via checkbox | Categorical |
| Washes hands after using toilets | The participant indicated that they wash their hands after using a toilet | Response by participant via checkbox | Categorical |
| Washes hands when dirty | The participant indicated that they wash their hands when their hands are dirty. | Response by participant via checkbox | Categorical |
| Washes hands before eating | The participant indicated that they wash their hands before eating. | Response by participant via checkbox | Categorical |
| **Water sources** | | | |
| Water Source: Surface Water | The participant indicated that their primary drinking water source includes surface water | Response by participant via checkbox | Categorical |
| Water Source: Rainwater | The participant indicated that their primary drinking water source includes rain water. | Response by participant via checkbox | Categorical |
| Water Source: Unprotected Spring/Well | The participant indicated that their primary drinking water source includes unprotected spring and/or an unrprotected well.   - 1- Participant answered “Yes” to using unprotected spring and/or unprotected well. - 0-Participant answered “No” to both using either an unprotected spring or unprotected well. | Response by participant via checkbox and collapsed by data inspector | Categorical |
| Water Source: Water tanker | The participant indicated that their primary drinking water source includes water tanker. | Response by participant via checkbox | Categorical |
| Water Source: Public Tap | The participant indicated that their primary drinking water source includes public taps. | Response by participant via checkbox | Cateogrical |
| Water Source: Tap in Courtyard | The participant indicated that their primary drinking water source includes a tap in courtyard (compound, yard or land) | Response by participant via checkbox | Categorical |
| Water Source: Tap in neighbour’s house | The participant indicated that their primary drinking water source includes the use of a tap from their neighbour’s house. | Response by participant via checkbox | Categorical |
| Water Source: Motorized borehole | The participant indicated that their primary drinking water source includes water from a motorized borehole | Response by participant via checkbox | Categorical |
| **Toilet infrastructures** | | | |
| Latrine Type: Flush Tank | The participant indicated that their primary toilet type they use is a Flush Tank. | Response by participant via checkbox | Categorical |
| Latrine Type: Hanging | The participant indicated that their primary toilet type they use is a Hanging toilet. | Response by participant via checkbox | Categorical |
| Latrine Type: Pit with no slab | The participant indicated that their primary toilet type they use is a Pit with no slab. | Response by participant via checkbox | Categorical |
| Latrine Type: Pit with slab | The participant indicated that their primary toilet type they use is a Pit with slab. | Response by participant via checkbox | Categorical |
| Latrine Type: Ventilated Pit | The participant indicated that their primary toilet type they use is a ventilated pit. | Response by participant via checkbox | Categorical |
| Latrine Type: Open defecation | The participant indicated that they openly defecate in bushes/fields as their primary toilet type. | Response by participant via checkbox | Categorical |
