## Supplementary material for "Risk factors influencing local cholera transmission in urban and rural endemic zones: the experience of Goma and Bukama, Democratic Republic of the Congo, 2021-2022.": Supp. S2

**Additional file S2:** Supplementary information on imputation details.

**Missing patterns in the different variables by study site.**

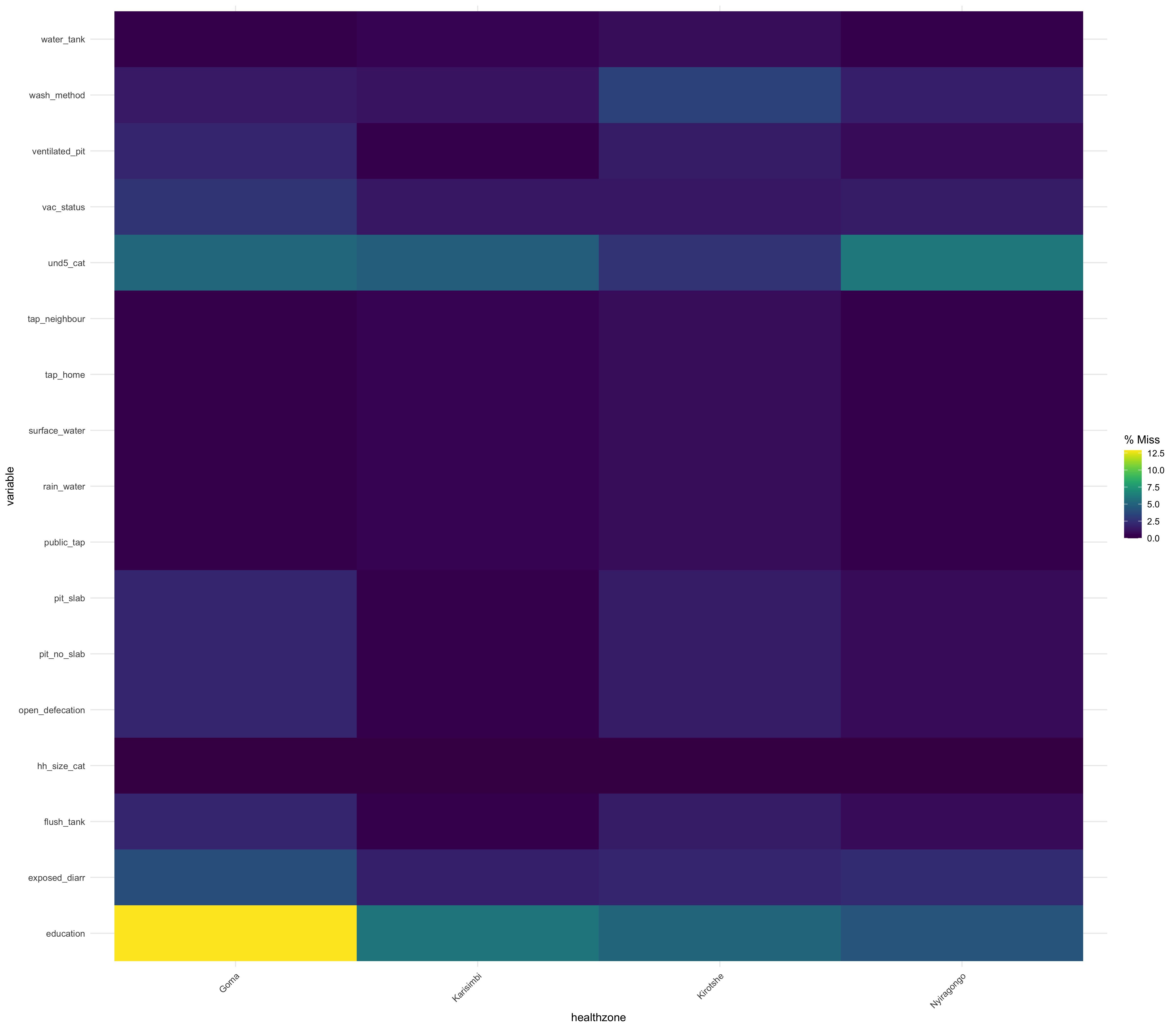

**Figure S2.1:** Missing data proportions in the predictor variables by Health Zone in Goma.

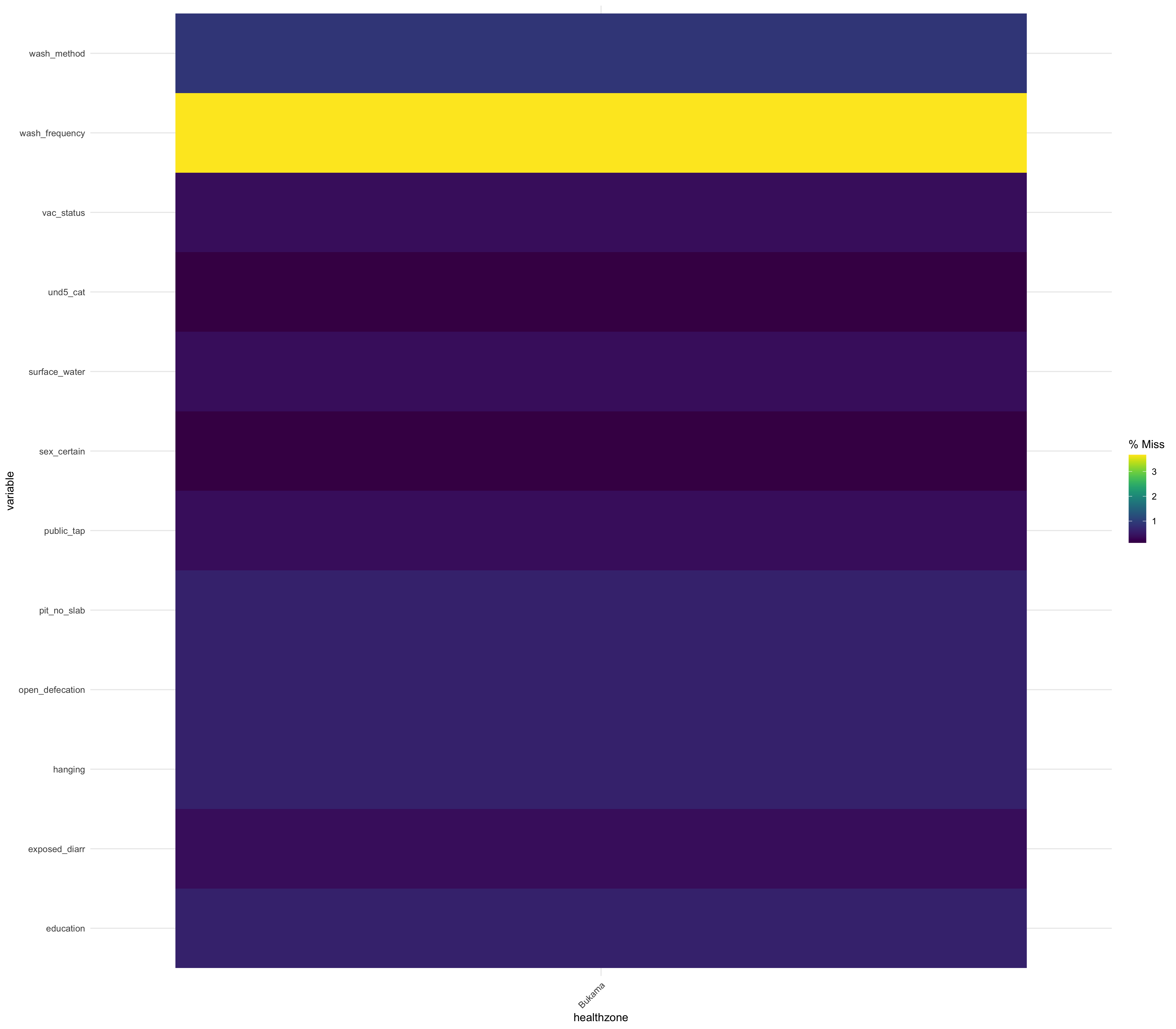

**Figure S2.2:** Missing data proportions in the predictor variables by Health Zone in Bukama.

**Imputation methodology.**

Imputation was conducted on variables for multivariate model-building purposes. The following variables were imputed utilizing the mice package in R.

**Table S2.1:** Study Site: **Goma**

| Variables | Method |
| --- | --- |
| Education, Under 5’s living in household, household size, age, hand washing method | “Polyreg” |
| Sex, OCV status, displacement status, washes hand before eating, washes hand before cooking, washes hands when dirty, exposed to suspected household case, flush tank, open defecation, surface water, public tap, rain water, tap in neighbour’s house, season, water tank, washes hand after toilet, pit with no slab, ventilated pit, tap in house | “Logreg” |

**Table S2.2:** Study Site: **Bukama**

| Variables | Method |
| --- | --- |
| Education, number of Under 5’s living in household, household size, age, hand washing method | “Polyreg” |
| Sex, OCV status, hand washing frequency, exposed to suspected household case, hanging toilet, unprotected spring, unprotected well, season, washes hands before cooking, washes hands after outside, open defecation, public tap, pit with no slab, surface water, motorized borehole | “Logreg” |

**Imputation Diagnostics: Strip Plots.**

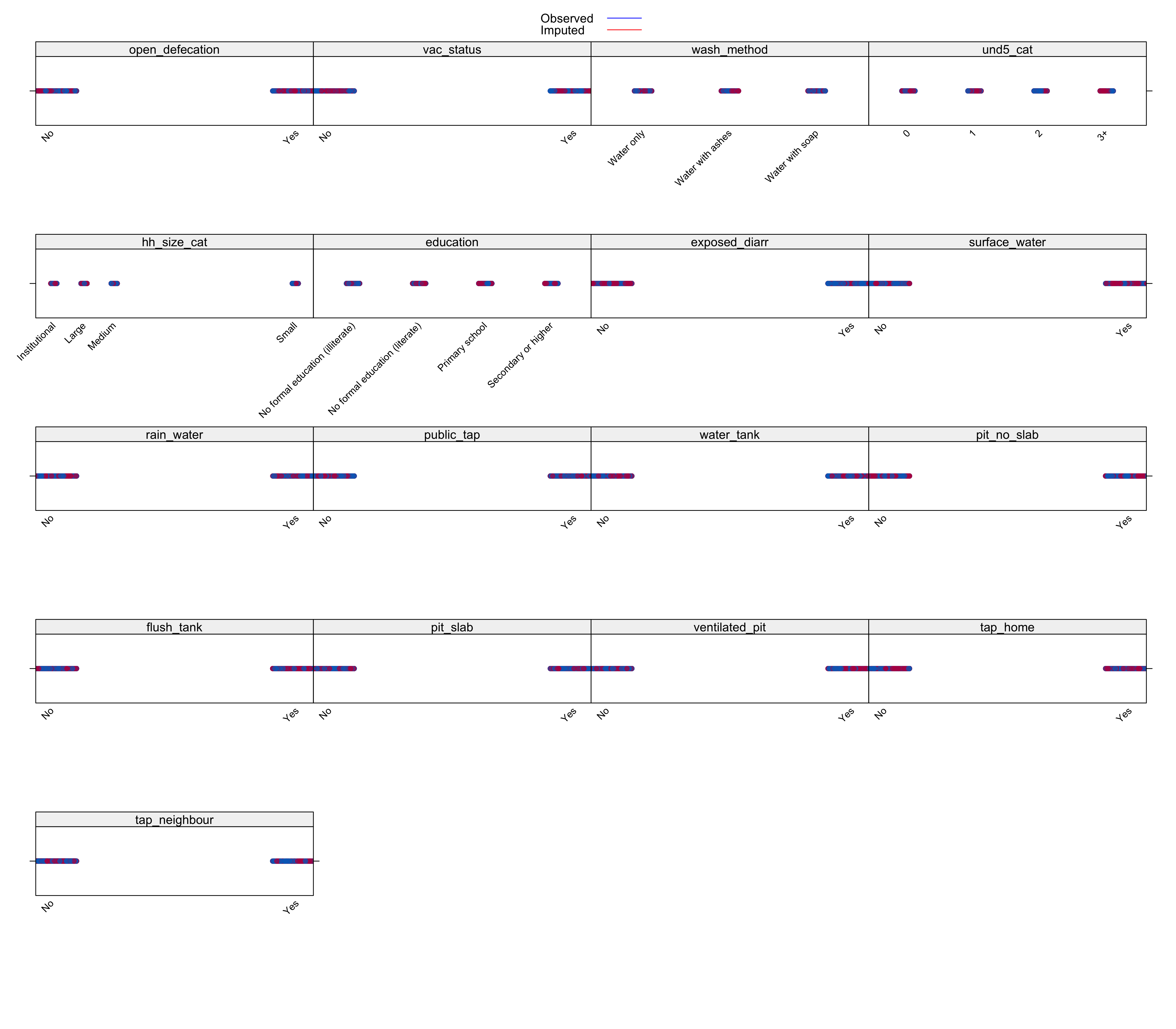

**Figure S2.3**: Strip plots for imputed and observed values of variables in Goma’s study population.

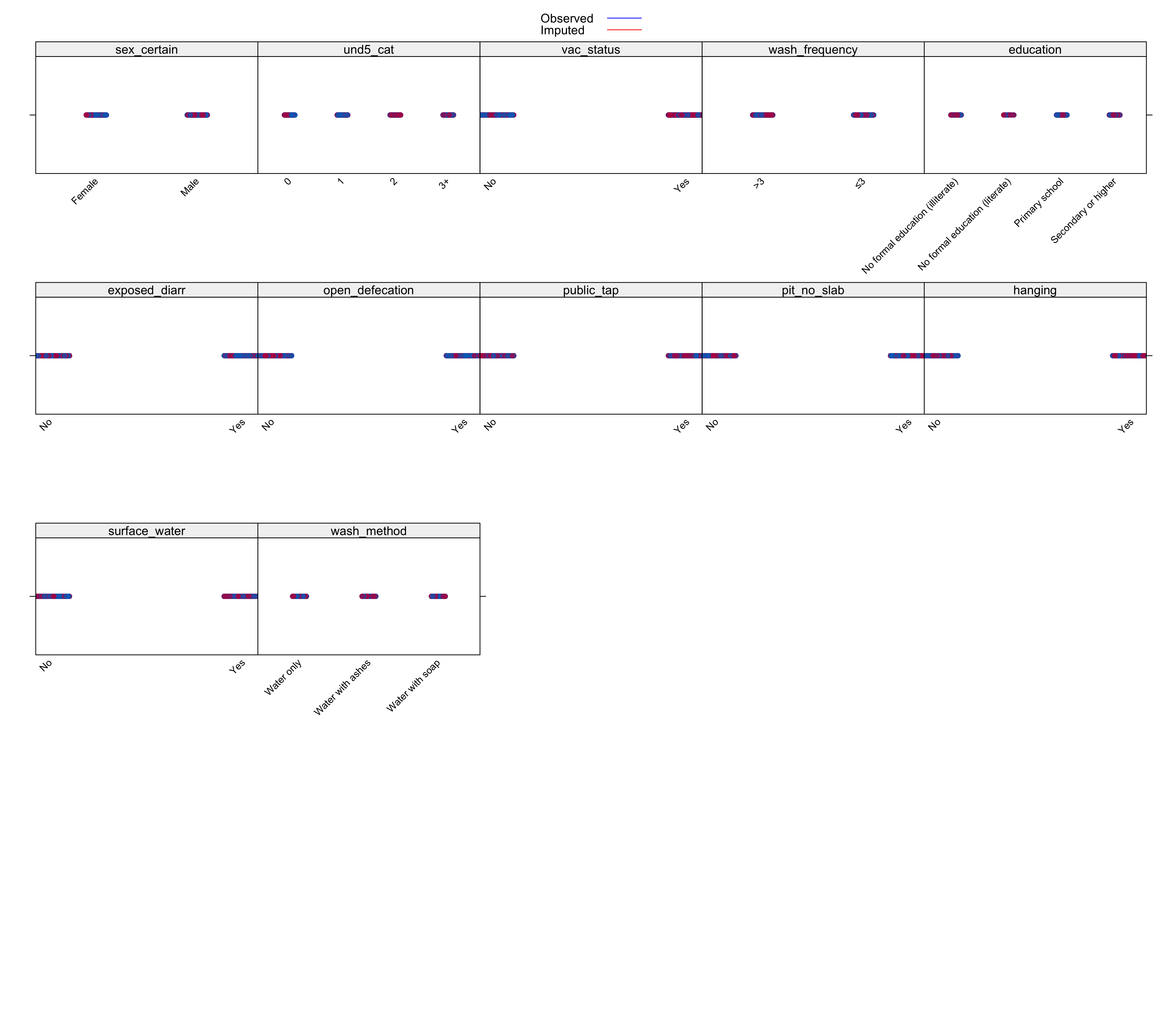

**Figure S2.4**: Strip plots for imputed and observed values of variables in Bukama’s study population

**Table S2.3.** Mixed-effect logistic regression model on non-imputed datasets for Goma and Bukama.

|  | **Goma** | | | |
| --- | --- | --- | --- | --- |
| **Characteristics** | **Negative**  n = 633 | **Positive**  n = 1,495 | **aOR (95% CI)** | **p-value** |
| **Sex, n (%)** |  |  |  |  |
| Female | 239 | 664 | -- | -- |
| Male | 394 | 831 | 1.19 (0.93, 1.51) | 0.16 |
| **Age Group, n (%)** |  |  |  |  |
| <5 | 183 | 383 | 0.84 (0.59, 1.20) | 0.34 |
| 5-14 | 61 | 552 | 3.30 (2.20, 4.94) | <0.001 |
| 15-24 | 145 | 228 | -- | -- |
| 25-44 | 173 | 256 | 0.69 (0.49, 0.98) | 0.04 |
| 45+ | 71 | 76 | 0.46 (0.29, 0.74) | 0.001 |
| **≥1 dose OCV, n (%)** |  |  |  |  |
| No | 518 | 1377 | -- | -- |
| Yes | 115 | 118 | 0.55 (0.38, 0.79) | 0.001 |
| **Displacement Status, n (%)** |  |  |  |  |
| No | 588 | 1,044 | -- | -- |
| Yes | 45 | 451 | 1.60 (0.89, 2.88) | 0.115 |
| **Education Level, n (%)** |  |  |  |  |
| No education | 112 | 442 | -- | -- |
| Primary school | 150 | 460 | 0.87 (0.63, 1.22) | 0.44 |
| Secondary or higher | 371 | 593 | 0.93 (0.66, 1.31) | 0.68 |
| **Season, n (%)** |  |  |  |  |
| Dry | 178 | 142 | -- | -- |
| Rainy | 455 | 1,353 | 3.89 (2.86, 5.28) | <0.001 |
| **Exposure to Cholera Cases, n (%)** |  |  |  |  |
| No | 423 | 1,101 | -- | -- |
| Yes | 210 | 394 | 1.77 (1.28, 2.45) | 0.001 |
| **Water Source: Surface Water, n (%)** |  |  |  |  |
| No | 564 | 1,235 | -- | -- |
| Yes | 69 | 260 | 2.07 (1.40, 3.08) | <0.001 |
| **Water Source: Rainwater, n (%)** |  |  |  |  |
| No | 611 | 1,469 | -- | -- |
| Yes | 22 | 26 | 0.51 (0.24, 1.08) | 0.08 |
| **Water Source: Water Tanker, n (%)** |  |  |  |  |
| No | 419 | 719 | -- | -- |
| Yes | 214 | 776 | 1.75 (1.22, 2.51) | 0.002 |
| **Handwashing Method, n (%)** |  |  |  |  |
| Water only | 244 | 479 | -- | -- |
| Water with ashes | 65 | 243 | 1.27 (0.83, 1.94) | 0.27 |
| Water with soap | 324 | 773 | 0.96 (0.71, 1.29) | 0.77 |
| **Toilet Type: Flush Tank, n (%)** |  |  |  |  |
| No | 471 | 1,451 | -- | -- |
| Yes | 162 | 44 | 0.42 (0.21, 0.83) | 0.01 |

|  | **Bukama** | | | |
| --- | --- | --- | --- | --- |
| **Variable** | **Negative**  n = 393 | **Positive**  n = 385 | **aOR (95% CI)** | **p-value** |
| **Sex, n (%)** |  |  |  |  |
| Female | 191 | 185 | -- | -- |
| Male | 202 | 200 | 1.04 (0.76, 1.44) | 0.81 |
| **Age Group, n (%)** |  |  |  |  |
| <5 | 36 | 40 | 0.89 (0.49, 1.62) | 0.71 |
| 5-14 | 68 | 79 | 1.16 (0.72, 1.88) | 0.54 |
| 15-24 | 127 | 98 | -- | -- |
| 25-44 | 119 | 126 | 1.31 (0.87, 1.98) | 0.20 |
| 45+ | 43 | 42 | 0.98 (0.55, 1.74) | 0.94 |
| **≥1 dose OCV, n (%)** |  |  |  |  |
| No | 52 | 104 | -- | -- |
| Yes | 341 | 281 | 0.59 (0.38, 0.92) | 0.02 |
| **Season, n (%)** |  |  |  |  |
| Dry | 153 | 87 | -- | -- |
| Rainy | 240 | 298 | 1.99 (1.39, 2.87) | <0.001 |
| **Water Source: Unprotected Spring/Well, n (%)** |  |  |  |  |
| No | 323 | 354 | -- | -- |
| Yes | 70 | 31 | 0.26 (0.15, 0.47) | <0.001 |
| **Toilet Type: Pit w/ no slab, n (%)** |  |  |  |  |
| No | 227 | 190 | -- | -- |
| Yes | 166 | 195 | 1.38 (0.83, 2.29) | 0.22 |
| **Toilet Type: Hanging Latrines over lake/river, n (%)** |  |  |  |  |
| No | 273 | 323 | -- | -- |
| Yes | 120 | 62 | 0.74 (0.42, 1.30) | 0.29 |
| **Exposure to Cholera Cases, n (%)** |  |  |  |  |
| No | 344 | 312 | -- | -- |
| Yes | 49 | 73 | 1.49 (0.91, 2.44) | 0.11 |
