## Supplementary material for "Risk factors influencing local cholera transmission in urban and rural endemic zones: the experience of Goma and Bukama, Democratic Republic of the Congo, 2021-2022.": Supp. S3

**Additional file S3:** Time and location description of suspected and laboratory-confirmed results distribution

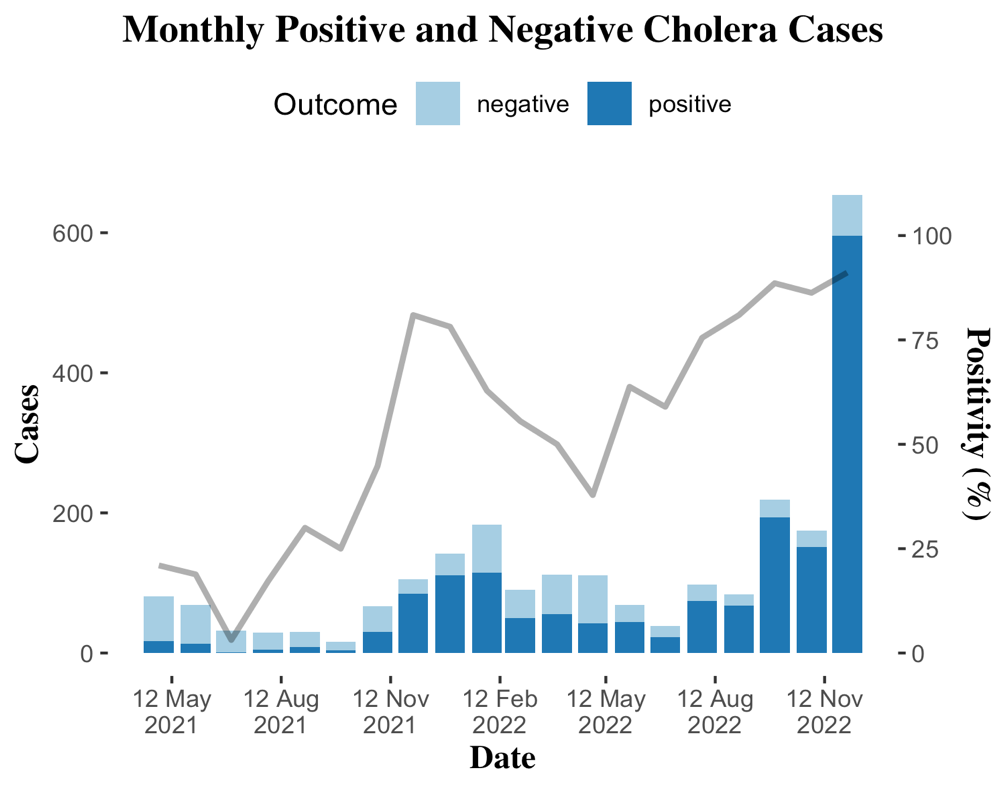

**A**

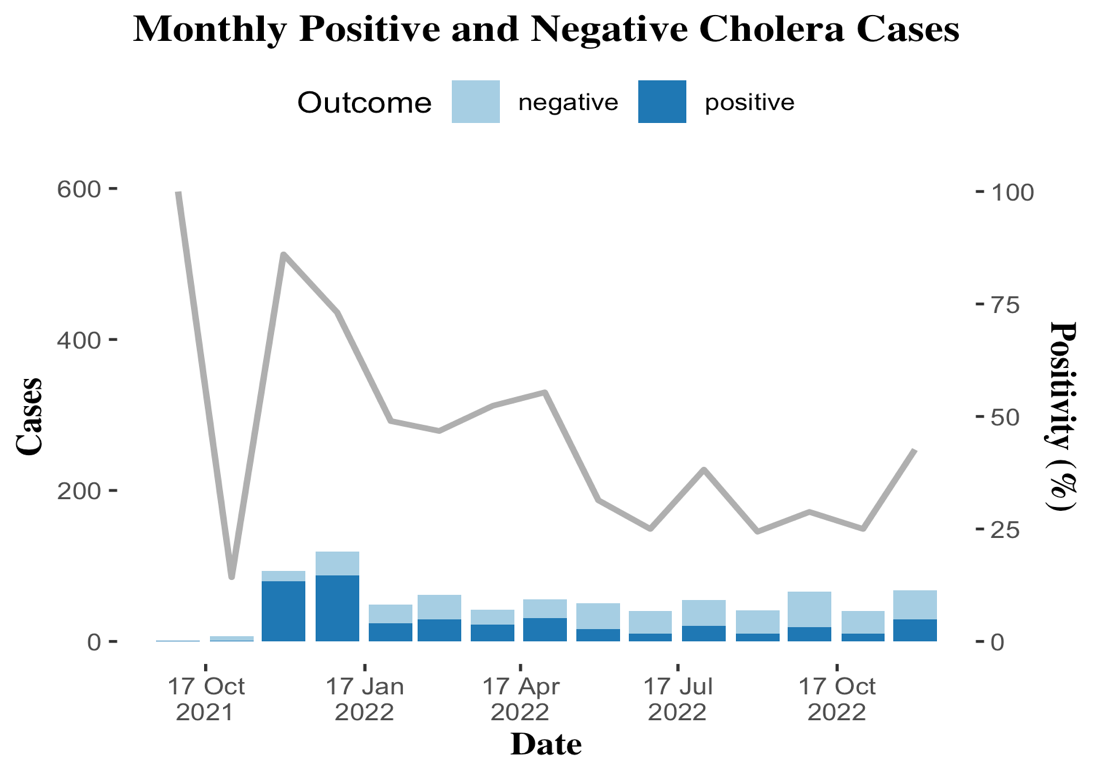

**B**

**Figure S3.1:** Monthly trends in overall positive and negative cholera cases in the study sites in the DRC between 2021 and 2022. A: Goma. B: Bukama

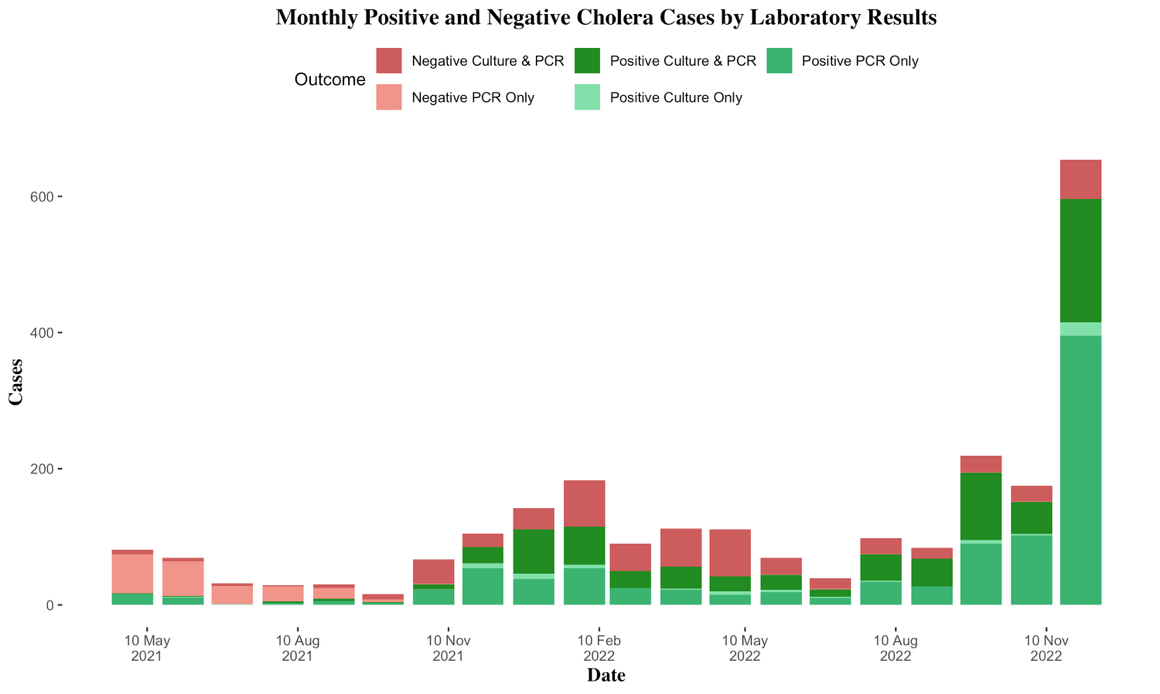

**A**

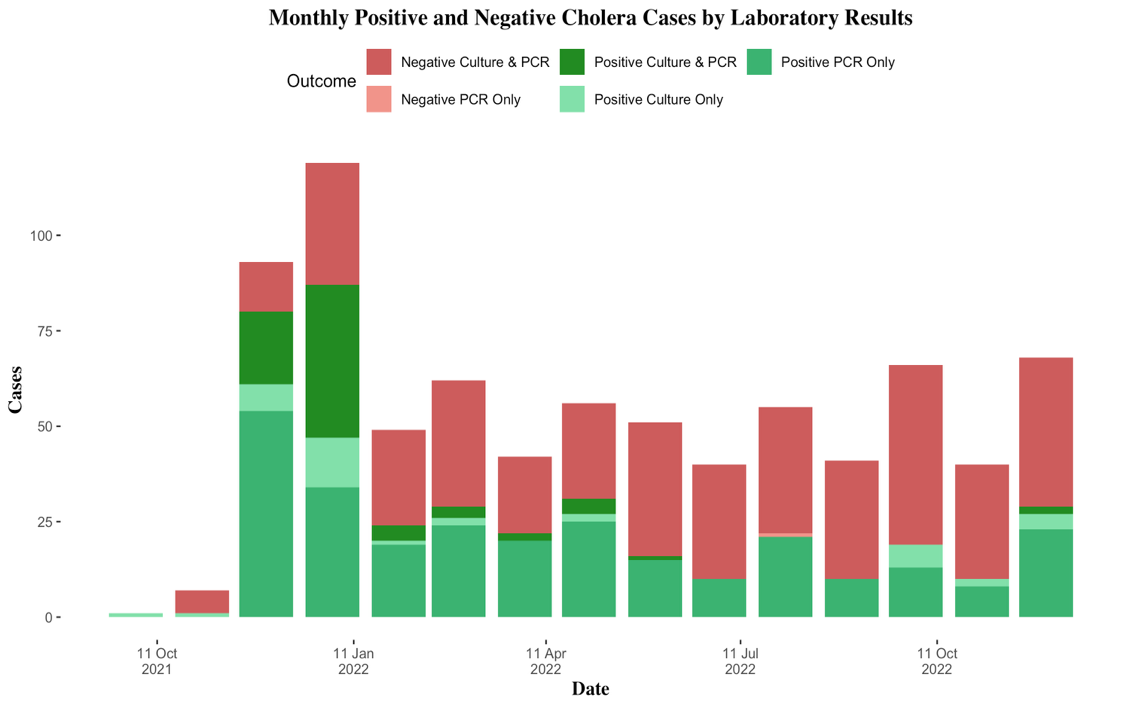

**B**

**Figure S3.2:** Monthly trends in positive and negative PCR/Culture cholera confirmations across the study sites in the DRC between 2021-2022. A: Goma. B: Bukama

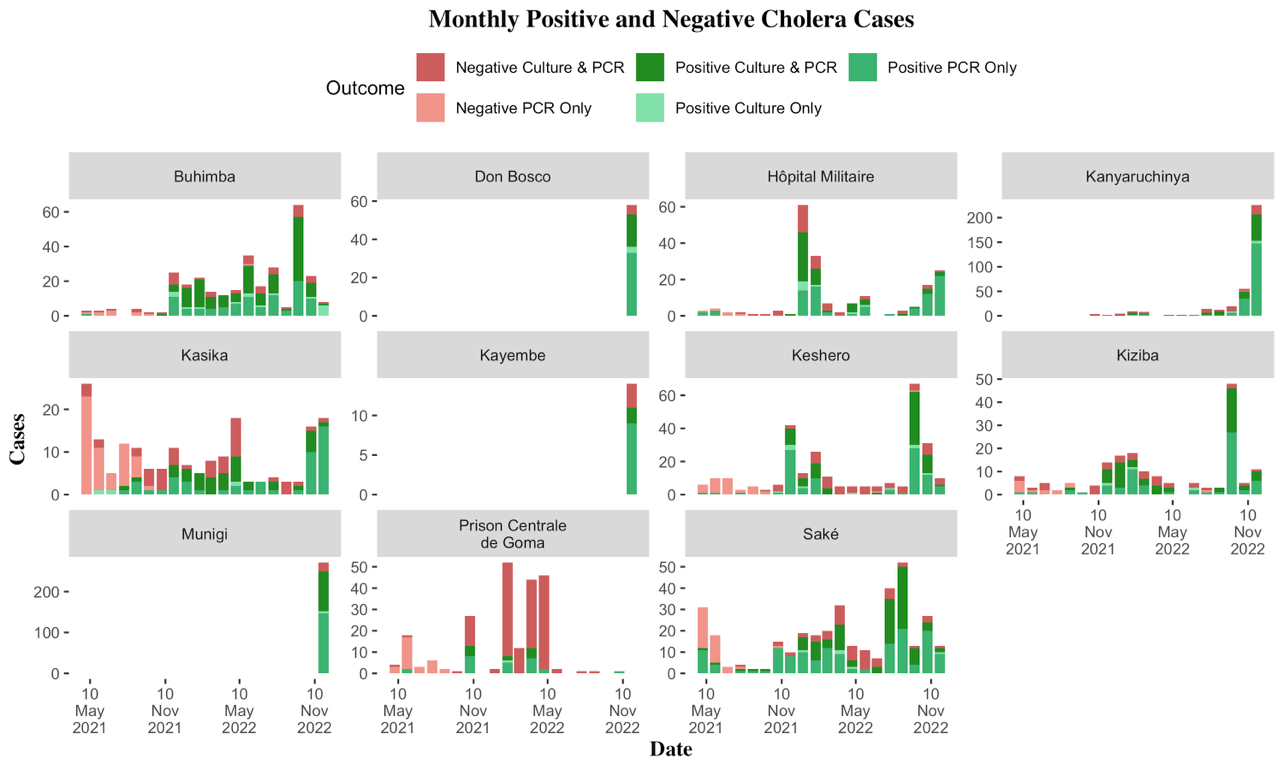

**Figure S3.3:** Monthly trends in positive and negative PCR/culture cholera confirmations across CTUs in Goma in the DRC between 2021 and 2022.

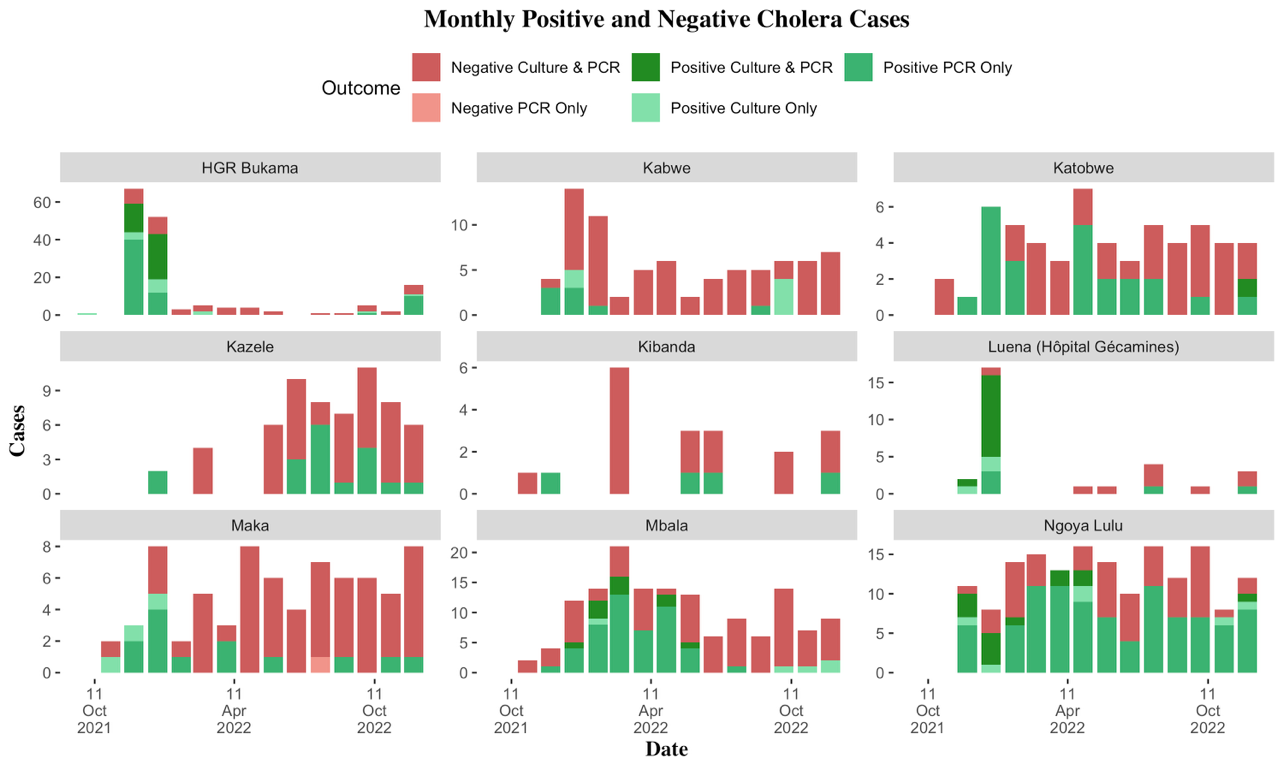

**Figure S3.4:** Monthly trends in positive and negative PCR/Culture cholera confirmations across CTUs in Goma in the DRC between 2021 and 2022.
