## Supplementary material for "Risk factors influencing local cholera transmission in urban and rural endemic zones: the experience of Goma and Bukama, Democratic Republic of the Congo, 2021-2022.": Supp. S4

**Additional file S4:** Univariate simple logistic regression output for laboratory-confirmed cholera positivity among suspected cases with Odds Ratios and 95% Confidence Intervals for Goma and Bukama, Democratic Republic of Congo, 2021–2022.

|  | **Goma** | | | **Bukama** | | |
| --- | --- | --- | --- | --- | --- | --- |
| **Characteristics** | **N** | **Crude OR (95% CI)** | **p-value** | **N** | **Crude OR (95% CI)** | **p-value** |
| **Sex, n (%)** | | | | | | |
| Female | 1,027 (43) | -- | -- | 380 (48) | -- | -- |
| Male | 1,378 (57) | 0.7429 (0.6207, 0.8879) | 0.0011 | 409 (52) | 1.0592 (0.8011, 1.4009) | 0.6864 |
| **Age Group, n (%)** | | | | | | |
| <5 | 640 (27) | 0.7174 (0.5548, 0.9280) | 0.0114 | 77 (9.7) | 1.4341 (0.8542, 2.4144) | 0.1727 |
| 5-14 | 706 (29) | 3.9365 (2.9537, 5.2944) | 0.0000 | 149 (19) | 1.5801 (1.0441, 2.3989) | 0.0309 |
| 15-24 | 418 (17) | -- | -- | 228 (29) | -- | -- |
| 25-44 | 476 (20) | 0.6861 (0.5357, 0.8784) | 0.0028 | 251 (32) | 1.4026 (0.9787, 2.0141) | 0.0659 |
| 45+ | 165 (6.9) | 0.4971 (0.3510, 0.7041) | 0.0001 | 85 (11) | 1.2957 (0.7855, 2.1379) | 0.3095 |
| **≥1 dose OCV, n (%)** | | | | | | |
| No | 2,111 (89) | -- | -- | 156 (20) | -- | -- |
| Yes | 254 (11) | 0.3729 (0.2863, 0.4856) | 0.0000 | 631 (80) | 0.4092 (0.2816, 0.5886) | 0.0000 |
| **Displacement Status, n (%)** | | | | | | |
| No | 1,870 (78) | -- | -- | 790 (100) |  |  |
| Yes | 535 (22) | 5.5120 (4.0891, 7.5900) | 0.0000 | 0 (0) |  |  |
| **Season, n (%)** | | | | | | |
| Dry | 379 (16) | -- | -- | 243 (31) | -- | -- |
| Rainy | 2,026 (84) | 3.7229 (2.9695, 4.6675) | 0.0000 | 547 (69) | 2.1712 (1.5939, 2.9715) | 0.0000 |
| **Exposure to suspect Cholera Cases in the household, n (%)** | | | | | | |
| No | 1,694 (72) | -- | -- | 664 (84) | -- | -- |
| Yes | 654 (28) | 0.7590 (0.6258, 0.9220) | 0.0053 | 123 (16) | 1.6078 (1.0908, 2.3866) | 0.0172 |
| **Education Level, n (%)** | | | | | | |
| No education | 590 (26) | -- | -- | 43 (5.5) | -- | -- |
| Primary school | 644 (29) | 0.7821 (0.5978,1.0210) | 0.0716 | 132 (17) | 0.4527 (0.2210,0.9074) | 0.0271 |
| Secondary or higher | 1,018 (45) | 0.4163 (0.3279,0.5257) | 0.0000 | 610 (78) | 0.6668 (0.3487,1.2442) | 0.2086 |
| **Children Under 5 in Household, n (%)** | | | | | | |
| 0 | 629 (28) | -- | -- | 122 (15) | -- | -- |
| 1 | 503 (22) | 1.8689 (1.4573,2.4034) | 0.0000 | 260 (33) | 1.4327 (0.9266,2.2301) | 0.1080 |
| 2 | 797 (35) | 2.5817 (2.0538,3.2520) | 0.0000 | 282 (36) | 1.8133 (1.1795,2.8091) | 0.0071 |
| 3+ | 358 (16) | 2.2778 (1.7132,3.0482) | 0.0000 | 125 (16) | 2.0309 (1.2263,3.3894) | 0.0062 |
| **Household Size, n (%)** | | | | | | |
| Institutional | 367 (16) | -- | -- | 29 (3.7) | -- | -- |
| Large | 1,021 (45) | 1.8526 (1.2961; 2.6285) | 0.0006 | 342 (43) | 2.9484 (1.9599,4.4855) | 0.0000 |
| Medium | 736 (32) | 1.7917 (1.2408, 2.5704) | 0.0017 | 279 (35) | 1.5879 (1.0426,2.4408) | 0.0328 |
| Small | 161 (7.0) | 0.4450 (0.3024, 0.6500) | 0.0000 | 140 (18) | 4.5411 (1.9678, 11.2270) | 0.0006 |
| **Handwashing Method, n (%)** | | | | | | |
| Water only | 782 (33) | -- | -- | 110 (14) | -- | -- |
| Water with ashes | 353 (15) | 1.7547 (1.3135,2.3634) | 0.0002 | 211 (27) | 0.8868 (0.5584,1.4065) | 0.6096 |
| Water with soap | 1,226 (52) | 1.1238 (0.9265,1.3620) | 0.2349 | 462 (59) | 0.8827 (0.5814,1.3384) | 0.5569 |
| **Washes Hands Before Cooking, n (%)** | | | | | | |
| No | 1,847 (77) | -- | -- | 635 (80) | -- | -- |
| Yes | 558 (23) | 1.4594 (1.1768,1.8190) | 0.0007 | 155 (20) | 0.7354 (0.5151,1.0462) | 0.0887 |
| **Washes Hands After Using Toilet, n (%)** | | | | | | |
| No | 1,162 (48) | -- | -- | 221 (28) | -- | -- |
| Yes | 1,243 (52) | 1.2159 (1.0209,1.4486) | 0.0285 | 569 (72) | 1.2577 (0.9214, 1.7196) | 0.1494 |
| **Washes Hands When Dirty, n (%)** | | | | | | |
| No | 1,259 (52) | -- | -- | 250 (32) | -- | -- |
| Yes | 1,146 (48) | 1.1888 (0.9977, 1.4173) | 0.0534 | 540 (68) | 1.1353 (0.8411,1.5338) | 0.4072 |
| **Washes Hands Before Eating, n (%)** | | | | | | |
| No | 121 (5.0) | -- | -- | 3 (0.4) |  |  |
| Yes | 2,284 (95) | 1.9016 (1.3086, 2.7494) | 0.0007 | 787 (100) |  |  |
| Water Source: **Surface Water, n (%)** | | | | | | |
| No | 2,008 (84) | -- | -- | 244 (31) | -- | -- |
| Yes | 385 (16) | 1.7278 (1.3351, 2.2585) | 0.0000 | 543 (69) | 0.8388 (0.6195, 1.1347) | 0.2545 |
| Water Source: **Rainwater, n (%)** | | | | | | |
| No | 2,341 (98) | -- | -- | 787 (100) |  |  |
| Yes | 52 (2.2) | 0.4162 (0.2392, 0.7243) | 0.0018 | 0 (0) |  |  |
| Water Source: **Water Tanker, n (%)** | | | | | | |
| No | 1,303 (54) | -- | -- | 787 (100) |  |  |
| Yes | 1,090 (46) | 2.0605 (1.7181, 2.4760) | 0.0000 | 0 (0) |  |  |
| Water Source: **Unprotected Spring, n (%)** | | | | | | |
| No | 2,388 (100) |  |  | 732 (93) | -- | -- |
| Yes | 5 (0.2) |  |  | 55 (7.0) | 0.6630 (0.3747, 1.1520) | 0.1493 |
| Water Source: **Unprotected Well, n (%)** | | | | | | |
| No | 2,391 (100) |  |  | 740 (94) | -- | -- |
| Yes | 2 (<0.1) |  |  | 47 (6.0) | 0.2574 (0.1196, 0.5059) | 0.0002 |
| Water Source: **Public Tap, n (%)** | | | | | | |
| No | 1,760 (74) | -- | -- | 618 (79) | -- | -- |
| Yes | 633 (26) | 0.5698 (0.4706, 0.6905) | 0.0000 | 169 (21) | 3.1773 (2.2110, 4.6250) | 0.0000 |
| Water Source: **Motorized Borehole, n (%)** | | | | | | |
| No | 2,393 (100) |  |  | 754 (96) | -- | -- |
| Yes | 0 (0) |  |  | 33 (4.2) | 0.3133 (0.1308,0.6742) | 0.0049 |
| Water Source: **Tap in Courtyard, n (%)** | | | | | | |
| No | 2,126 (89) | -- | -- | 784 (100) |  |  |
| Yes | 267 (11) | 0.4430 (0.3421,0.5741) | 0.0000 | 3 (0.4) |  |  |
| Water Source: **Tap in Neighbour's House, n (%)** | | | | | | |
| No | 2,179 (91) | -- | -- | 784 (100) |  |  |
| Yes | 214 (8.9) | 0.5649 (0.4244,0.7545) | 0.0001 | 3 (0.4) |  |  |
| Toilet Type: **Pit Latrine Without Slab, n (%)** | | | | | | |
| No | 902 (38) | -- | -- | 421 (54) | -- | -- |
| Yes | 1,479 (62) | 3.0431 (2.5389, 3.6515) | 0.0000 | 364 (46) | 1.3915 (1.0505, 1.8451) | 0.0214 |
| Toilet Type: **Hanging Toilet, n (%)** | | | | | | |
| No | 2,377 (100) |  |  | 602 (77) | -- | -- |
| Yes | 4 (0.2) |  |  | 183 (23) | 0.4338 (0.3057, 0.6107) | 0.0000 |
| Toilet Type: **Pit Latrine With Slab, n (%)** | | | | | | |
| No | 1,845 (77) | -- | -- | 747 (95) | -- | -- |
| Yes | 536 (23) | 0.7128 (0.5817,0.850) | 0.0011 | 38 (4.8) | 1.1433 (0.5948, 2.2167) | 0.6856 |
| Toilet Type: **Ventilated Pit, n (%)** | | | | | | |
| No | 2,328 (98) | -- | -- | 776 (99) |  |  |
| Yes | 53 (2.2) | 2.4236 (1.2015, 5.5769) | 0.0219 | 9 (1.1) |  |  |
| Toilet Type: **Open Defecation, n (%)** | | | | | | |
| No | 2,324 (98) | -- | -- | 602 (77) | -- | -- |
| Yes | 57 (2.4) | 2.20217 (1.0601, 4.2630) | 0.0450 | 183 (23) | 1.3519 (0.9704, 1.8878) | 0.0754 |
| Toilet Type: **Flush Tank, n (%)** | | | | | | |
| No | 2,151 (90) | -- | -- | 776 (99) |  |  |
| Yes | 230 (9.7) | 0.0881 (0.0627, 0.1216) | 0.0000 | 9 (1.1) |  |  |
