## Supplementary material for "Risk factors influencing local cholera transmission in urban and rural endemic zones: the experience of Goma and Bukama, Democratic Republic of the Congo, 2021-2022.": Supp. S5

**Additional file S5:** Univariate and multivariate simple logistic regression (**A.** only CTU HGR Bukama) and mixed-effect logistic regression (**B.** all CTUs except HGR Bukama) outputs for laboratory-confirmed cholera positivity among suspected cases with Odds Ratios and 95% Confidence Intervals, Bukama, Democratic Republic of Congo, 2021–2022

|  |  | | **Bukama – only CTU HGR Bukama** | | | |
| --- | --- | --- | --- | --- | --- | --- |
| **Variable** | **Negative**  n = 45 | **Positive**  n = 118 | | **OR (95% CI)** | **aOR (95% CI)** | **p-value** |
| **Sex, n (%)** |  |  | |  |  |  |
| Female | 19 | 57 | | -- | -- | -- |
| Male | 26 | 61 | | 0.78 (0.39, 1.56) | 1.04 (0.45, 2.39) | 0.74 |
| **Age Group, n (%)** |  |  | |  |  |  |
| <5 | 8 | 23 | | 1.25 (0.42, 3.74) | 1.31 (0.36, 4.76) | 0.9 |
| 5-14 | 10 | 40 | | 1.74 (0.63, 4.80) | 1.47 (0.45, 4.77) | 0.38 |
| 15-24 | 10 | 23 | | -- | -- | -- |
| 25-44 | 11 | 20 | | 0.79 (0.28, 2.25) | 0.60 (0.17, 2.12) | 0.36 |
| 45+ | 6 | 12 | | 0.87 (0.25, 2.97) | 0.69 (0.16, 3.02) | 0.26 |
| **≥1 dose OCV, n (%)** |  |  | |  |  |  |
| No | 15 | 50 | | -- | -- | -- |
| Yes | 30 | 68 | | 0.68 (0.33, 1.40) | 0.57 (0.22, 1.48) | 0.66 |
| **Season, n (%)** |  |  | |  |  |  |
| Dry | 8 | 0 | | -- | -- | -- |
| Rainy | 37 | 118 | | 2.93 (1.38, Inf)* | 3e+17 (0, Inf) | 0.99 |
| **Water Source: Unprotected Spring/Well, n (%)** |  |  | |  |  |  |
| No | 20 | 98 | | -- | -- | -- |
| Yes | 25 | 20 | | 0.16 (0.08, 0.35) | 0.17 (0.07, 0.43) | <0.001 |
| **Toilet Type: Pit w/ no slab, n (%)** |  |  | |  |  |  |
| No | 5 | 9 | | -- | -- | -- |
| Yes | 40 | 109 | | 1.51 (0.48, 4.79) | 1.85 (0.41, 8.36) | 0.27 |
| **Toilet Type: Hanging Latrines over lake/river, n (%)** |  |  | |  |  |  |
| No | 44 | 115 | | -- | -- | -- |
| Yes | 1 | 3 | | 1.15 (0.12, 11.33) | 3.12 (0.20, 49.00) | 0.96 |
| **Exposure to suspect Cholera Cases in the household, n (%)** |  |  | |  |  |  |
| No | 33 | 64 | | -- | -- | -- |
| Yes | 12 | 54 | | 2.32 (1.09, 4.93) | 3.15 (1.21, 8.19) | 0.56 |

**Table S5.1:** Multivariate simple logistic regression results for **CTU HGR Bukama.**

* Univariate results obtained with exact logistic regression – relevant technique for small sample sizes with extreme imbalance in data (“quasi-complete separation”) like in the “Season” variable**Table S5.2:** Description of water users in **CTU HGR Bukama**

N = **163**

Surface water users = 29/N (**18%**) – positivity rates among users = 23/29 (79%)

Unprotected source water users = 45/N (**28%**) – positivity rates among users = 20/45 (44%)

Public tap water users = 156/N (**96%**) – positivity rates among users = 113/156 (69%)

Overlap between water sources =

|  | Unprotected source NO | Unprotected source YES |
| --- | --- | --- |
| Surface water NO | **96** | **38** |
| Surface water YES | **22** | **7** |

|  | Public tap NO | Public tap YES |
| --- | --- | --- |
| Surface water NO | **4** | **130** |
| Surface water YES | **3** | **26** |

|  | Unprotected source NO | Unprotected source YES |
| --- | --- | --- |
| Public tap NO | **2** | **5** |
| Public tap YES | **116** | **40** |

Among unprotected source users, 15% (7/45) also drink surface water and 89% (40/45) also drink public tap water.

|  |  | | **Bukama – all CTUs except HGR Bukama** | | | |
| --- | --- | --- | --- | --- | --- | --- |
| **Variable** | **Negative**  n = 355 | **Positive**  n = 272 | | **OR (95% CI)** | **aOR (95% CI)** | **p-value** |
| **Sex, n (%)** |  |  | |  |  |  |
| Female | 177 | 128 | | -- | -- | -- |
| Male | 178 | 144 | | 1.01 (0.72, 1.42) | 1.08 (0.76, 1.54) | 0.67 |
| **Age Group, n (%)** |  |  | |  |  |  |
| <5 | 29 | 17 | | 0.82 (0.40, 1.69) | 0.69 (0.33, 1.46) | 0.33 |
| 5-14 | 58 | 41 | | 1.12 (0.66, 1.91) | 1.12 (0.65, 1.94) | 0.67 |
| 15-24 | 120 | 75 | | -- | -- | -- |
| 25-44 | 111 | 109 | | 1.39 (0.91, 2.11) | 1.50 (0.97, 2.31) | 0.07 |
| 45+ | 37 | 30 | | 1.18 (0.64, 2.20) | 1.04 (0.55, 1.95) | 0.91 |
| **≥1 dose OCV, n (%)** |  |  | |  |  |  |
| No | 37 | 55 | | -- | -- | -- |
| Yes | 318 | 217 | | 0.56 (0.33, 0.96) | 0.48 (0.28, 0.84) | 0.009 |
| **Season, n (%)** |  |  | |  |  |  |
| Dry | 147 | 88 | | -- | -- | -- |
| Rainy | 208 | 184 | | 1.61 (1.12, 2.30) | 1.76 (1.22, 2.55) | 0.004 |
| **Water Source: Unprotected Spring/Well, n (%)** |  |  | |  |  |  |
| No | 310 | 260 | | -- | -- | -- |
| Yes | 45 | 12 | | 0.56 (0.25, 1.27) | 0.46 (0.20, 1.05) | 0.06 |
| **Toilet Type: Pit w/ no slab, n (%)** |  |  | |  |  |  |
| No | 277 | 185 | | -- | -- | -- |
| Yes | 128 | 87 | | 1.34 (0.86, 2.08) | 1.15 (0.66, 2.00) | 0.61 |
| **Toilet Type: Hanging Latrines over lake/river, n (%)** |  |  | |  |  |  |
| No | 233 | 212 | | -- | -- | -- |
| Yes | 122 | 60 | | 0.75 (0.47, 1.19) | 0.68 (0.37, 1.23) | 0.20 |
| **Exposure to suspect Cholera Cases in the household, n (%)** |  |  | |  |  |  |
| No | 317 | 253 | | -- | -- | -- |
| Yes | 38 | 19 | | 0.87 (0.46, 1.63) | 0.80 (0.42, 1.53) | 0.51 |

**Table S5.3:** Mixed-effect logistic regression results for **all CTUs except HGR Bukama**

**Table S5.4:** Description of water users in **all** **CTUs except HGR Bukama**

N = **627**

Surface water users = 517/N (**82%**) – positivity rates among users = 239/517 (46%)

Unprotected source water users = 57/N (**9%**) – positivity rates among users = 12/57 (21%)

Public tap water users = 14/N (**2%**) – positivity rates among users = 7/14 (50%)

Overlap between water sources =

|  | Unprotected source NO | Unprotected source YES |
| --- | --- | --- |
| Surface water NO | **55** | **55** |
| Surface water YES | **515** | **2** |

|  | Public tap NO | Public tap YES |
| --- | --- | --- |
| Surface water NO | **97** | **13** |
| Surface water YES | **516** | **1** |

|  | Unprotected source NO | Unprotected source YES |
| --- | --- | --- |
| Public tap NO | **556** | **57** |
| Public tap YES | **14** | **0** |

Among unprotected source users, 3% (2/57) also drink surface water and 0% (0/57) also drink public tap water.
